# Generalizability of proteomic risk prediction across biobanks reveals dependence on phenotype definitions

**DOI:** 10.64898/2026.10.01.26364038

**Authors:** Sophie J. Parsa, Yon Ho Jee, M. Austin Argentieri, Jeremy Guez, Wenhan Lu, Evin M. Padhi, Elizabeth Kiernan, Reza Jabal, Mitja I. Kurki, FinnGen, Benjamin M. Neale, Lee Lichtenstein, Michael E. Talkowski, Stephen B. Montgomery, Niall J. Lennon, Alicia R. Martin, Mark J. Daly, Konrad J. Karczewski

## Abstract

Advances in high-throughput proteomics technologies have enabled the assessment of dynamic health states across biobank-scale cohorts. Disease prediction models built on these data have higher accuracy than baseline clinical models for a broad range of diseases, and provide avenues to understand disease pathogenesis. However, the generalizability of these prediction models across cohorts has yet to be established at scale. Here, we train models for 15 diseases in the UK Biobank (UKB; n=53,026) based on Olink proteomics data, and find high accuracy for disease prediction (mean AUC=0.74; range 0.56-0.89). These models have improved accuracy over models using only clinical factors (mean ΔAUC = 0.03), and do not depend on model architecture, with simpler models (e.g. L2) performing as well as complex models (e.g. transformer). We assess generalizability of the UKB-trained models in two external cohorts: FinnGen (FG; n=5,865) and *All of Us* (AoU; n=7,405), spanning two Olink platforms. We find that proteomics-based prevalent (classification) and incident (prediction over next 5 years) disease models largely generalize across cohorts, but performance varies across diseases. Specifically, 10 of 14 prevalent and 13 of 15 incident disease models show no significant decrease in performance across any biobank. Adjusting for demographic, ancestry, and technical covariates, we demonstrate that differences in phenotyping quality are likely the major drivers of variability across cohorts. These results indicate that proteomic risk models can be powerful and generalizable predictors of disease across multiple cohorts.

## Introduction

Early disease detection improves clinical outcomes for a broad range of diseases including chronic respiratory disease (*1*), reproductive system diseases (*2*), cancer (*3*), and cardiovascular diseases. Some of the greatest efforts in early detection have focused on cancer, leading to substantial reductions in mortality for several malignancies: early screening for prostate specific antigen (PSA) can reduce mortality by 20%, although overdiagnosis is a significant concern (*4*). Furthermore, the five year survival rate for breast cancer has increased from 75% in 1975 to 90% in 2011, and this mortality reduction has largely been attributed to early detection and screening through mammograms (*5*).

Further, accurate biomarkers for disease are essential for disease management. In some cases, a small number of key risk factors can provide profound insight into disease pathogenesis, such as in the case of hypertension and high cholesterol for coronary artery disease (*6*). However, for many diseases, most of the predictive signal comes from biological readouts that reflect risk from other exogenous sources like environmental influences and behavior. These may reflect disease processes that are already underway before diagnosis or coincidental (non-causal) factors relevant to disease: such readouts are valuable not only for prediction but also as integral components of drug development pipelines, including the development of surrogate endpoints.

Proteomics may provide an avenue for early detection of multiple human diseases in a single assay. The proteome encodes not only elements of the genome through regulation of protein expression with certain variants, but also non-genetically driven biological processes that contribute to disease — including demographic characteristics, such as age and sex, and lifelong accumulation of environmental exposures, such as smoking (*7*), which alter protein expression (*8*, *9*). This rich information makes proteomics well-suited for disease prediction: within a single assay, some proteins may represent novel, druggable targets while others serve as readouts of general disease processes such as inflammation or disease-related behaviors. Since biobank-scale proteomics characterizes expression of thousands of proteins, it has the potential to lead to the development of screening tools while identifying both causal risk factors and non-causal biomarkers.

Prevalent disease classification and incident disease prediction models derived from proteomic data can be applied to predict hundreds of disease endpoints, achieving high performance for a broad range of diseases, particularly those related to diabetes (*10*). Furthermore, the choice of models has been shown to affect performance, as some recent analyses showed that neural networks outperformed linear models for some outcomes or with the addition of phenotypic correlation data, and for all outcomes the addition of proteomics to clinical models improved prediction accuracy (*11*, *12*). Importantly, proteomics models built in UK Biobank (UKB) have been shown to improve disease risk prediction compared to both standard clinical models and models developed using other data modalities, which have been validated in small cohorts (*13*). Even for diseases that are strongly predicted by clinical lab measurements, proteomics can still improve risk prediction; for instance, a protein score for type 2 diabetes outperforms a polygenic risk score and glycated haemoglobin (HbA1c), which is a well established clinical marker for type 2 diabetes (*14*). This improvement of proteomics based risk prediction over polygenic risk score has been shown across a wide range of diseases (*15*). However, for proteomics to serve as a clinically significant screening tool or biomarker representing underlying disease biology, the signal must be applicable across diverse cohorts, including different healthcare systems and separate proteomic batches, and this transferability has not yet been systematically assessed.

Here, we assess generalizability of proteomic models by building models to classify prevalent and predict incident diseases in the UKB, with comparison to models built on clinical lab values (e.g. blood cell counts, liver function tests, HbA1c, and creatinine). We train a series of models in the UKB and then evaluate in two external cohorts: FinnGen (FG) and *All of Us* (AoU), using multiple model architectures, and find that these models largely generalize across cohorts, healthcare systems, demographic axes (e.g. age and sex), and other participant characteristics such as genetic ancestry and BMI. Finally, we explore the importance of phenotyping and other factors that may influence performance of proteomics based disease prediction models across cohorts, as well as the specificity of these models.

## Results

We used proteomic data available from UKB (n=53,206; Olink Explore 3072), AoU (n=7,405; Olink Explore HT; the generation of which is described in a forthcoming flagship manuscript [Padhi et al., 2026]) and two cohorts in FinnGen (n=5,865, including 1,925 Olink Explore HT and 3,940 Olink Explore 3072). Across cohorts, participants are predominantly middle- to older-aged adults at enrollment, with differences in demographic composition (e.g., greater geographic diversity in AoU). Summary statistics for age at proteomics sample collection, sex, baseline body mass index (BMI), and smoking status for the proteomics cohort in each biobank are shown in Table 1, along with disease prevalence at baseline and five year incident case counts during follow-up.

**Table 1.** Cohort characteristics. We consider three biobanks in our analysis: the UK Biobank, FinnGen, and AoU. Participant characteristics are calculated for participants at available baseline enrollment dates. Smoking status in UKB and FinnGen is determined using self reported smoking status (current or previous vs never) and in AoU based on ≥100 lifetime cigarettes. For all cohorts, prevalent and 5-year incident case counts are computed from ICD9 and ICD10 codes. For UKB, baseline medication, lab, and interview codes are also included, and for FinnGen, Risteys endpoints, which integrate insurance reimbursement and medication codes, are also included (Tables S1-2). Prevalent cases are excluded from incident analysis.

| Participant characteristics | UKB | AoU | FinnGen |  |  |  |
| --- | --- | --- | --- | --- | --- | --- |
| Age, median [range] | 59 [39–71] | 51 [18–97] | 55 [16–92] |  |  |  |
| Male sex, % | 46.1% | 45.9% | 47.8% |  |  |  |
| BMI, median [range] | 26.8<br>[14.3–69.0] | 27.4<br>[14.7–81.5] | 26.2<br>[14.2–56.1] |  |  |  |
| Ever smoker, % | 45.7% | 33.8% | 45.4% |  |  |  |
| Disease | Prevalent |  |  | Incident |  |  |
|  | UKB | AoU | FinnGen | UKB | AoU | FinnGen |
| Asthma | 7,890 (14.9%) | 1,007 (13.6%) | 479 (8.2%) | 1039 (2.3%) | 232 (3.6%) | 81 (1.5%) |
| Osteoarthritis | 7,574 (14.3%) | 1,484 (20.0%) | 864 (14.7%) | 2566 (5.6%) | 472 (8.0%) | 194 (3.9%) |
| Ischemic heart disease | 6,191 (11.7%) | 916 (12.4%) | 410 (7.0%) | 2271 (4.8%) | 349 (5.4%) | 160 (2.9%) |
| Type 2 diabetes | 2,958 (5.6%) | 1,285 (17.4%) | 582 (9.9%) | 831 (1.7%) | 300 (4.9%) | 207 (3.9%) |
| Kidney disease | 1,143 (2.2%) | 466 (6.3%) | 47 (0.8%) | 1051 (2.0%) | 241 (3.5%) | 81 (1.4%) |
| Rheumatoid arthritis | 1,128 (2.1%) | 198 (2.7%) | 108 (1.8%) | 412 (0.79%) | 51 (0.7%) | 128 (2.2%) |
| All stroke | 1,075 (2.0%) | 230 (3.1%) | 152 (2.6%) | 509 (0.98%) | 105 (1.5%) | 96 (1.7%) |
| COPD | 806 (1.5%) | 327 (4.4%) | 95 (1.6%) | 857 (1.6%) | 134 (1.9%) | 49 (0.8%) |
| Osteoporosis | 1,303 (2.5%) | 482 (6.5%) | 63 (1.1%) | 592 (1.1%) | 152 (2.2%) | 53 (0.9%) |
| Ischemic stroke | 484 (0.91%) | 208 (2.8%) | 134 (2.3%) | 454 (0.86%) | 88 (1.2%) | 75 (1.3%) |
| Ulcerative colitis | 507 (0.96%) | 83 (1.1%) | 40 (0.7%) | 135 (0.26%) | <20 (0.3%) | 109 (1.9%) |
| Macular degeneration | 320 (0.60%) | 289 (3.9%) | 185 (3.2%) | 308 (0.58%) | 107 (1.5%) | 127 (2.2%) |
| Liver disease | 393 (0.74%) | 488 (6.6%) | 42 (0.7%) | 325 (0.62%) | 244 (3.5%) | 28 (0.5%) |
| Parkinson's disease | 215 (0.41%) | 29 (0.4%) | 329 (5.6%) | 461 (0.87%) | 27 (0.4%) | 36 (0.7%) |
| Alzheimer's disease | n/a | n/a | n/a | 152 (0.29%) | <20 (0.3%) | 119 (2.0%) |

We selected 15 major chronic diseases with at least 100 incident cases in the UKB for model development. We built models of each disease using protein expression values as input (Fig. 1A), using several architectures: L1 (Lasso) and L2 (ridge) penalized logistic regressions, light gradient boosting machines (LGBMs), multilayer perceptrons (MLPs), and a transformer (fine-tuned for each disease). We trained using 90% of the UKB (n=47,723), and evaluated in 10% holdout UKB (n=5,303), as well as FinnGen and AoU. In the case of prevalent disease, the model classifies whether or not someone currently has the disease at the time of blood collection for proteomics, and in the case of incident disease, the model predicts whether the person will develop the disease in the next five years and excludes those with prevalent disease at baseline.

**Figure 1.**
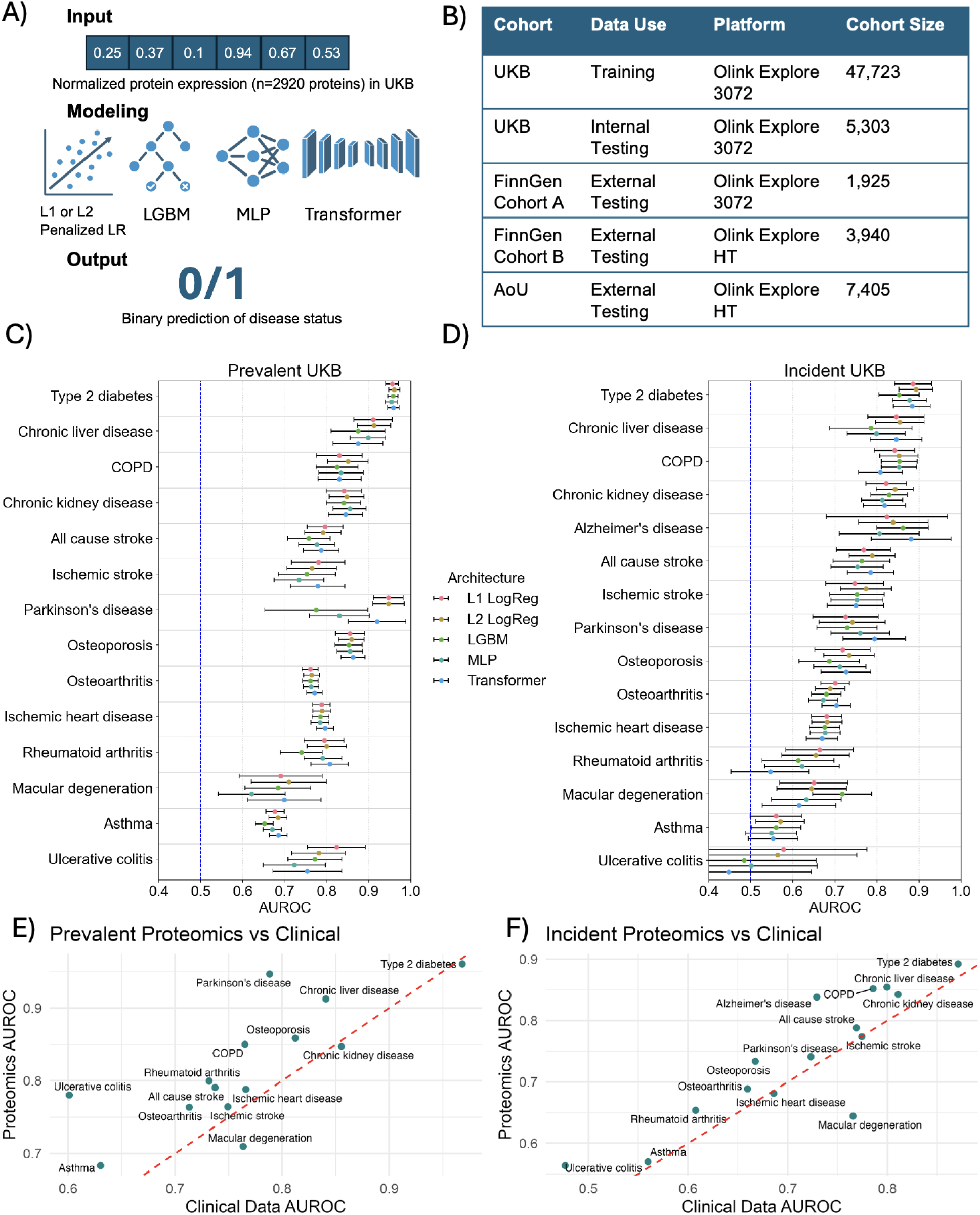
Multi-biobank proteomic study design evaluates model architectures for disease prediction. A) Disease models are trained in the UKB for prevalent and incident disease based on protein expression values, using five architectures: L1 penalized (LASSO) logistic regression, L2 penalized (ridge) logistic regression, light gradient boosting machines (LGBM), multilayer perceptron (MLP), and a fine tuned transformer. B) Models are trained using UKB, internally evaluated in UKB hold out data, and externally evaluated in FinnGen and AoU. FinnGen cohort A and UKB share the same Olink panel while FinnGen cohort B and AoU have a newer, larger panel. C-D) Performance in UKB holdout data of prevalent classification (C) and 5 year incident prediction (D) models. E-F) Using the same training testing split as proteomics models, we train models using baseline clinical data using ridge regression and assess in hold out test data. We find increased average AUROC for proteomics compared to clinical data for prevalent classification (t = 3.16, p = 0.0075) and 5 year incident prediction disease (t = 2.12, p = 0.052).

For both prevalent and five year incident diseases, our models showed strong performance in UKB holdout data. At least one architecture obtained an AUROC above 0.7 for 13 out of the 14 prevalent disease models (Fig. 1C) and 11 out of the 15 incident disease models (Fig. 1D). There were no consistent differences in performance across architectures in the UKB, with linear and non-linear models achieving similar performances in holdout data. As a comparator, we also trained disease prediction models based on a comprehensive set of clinical data, including age, sex, complete blood count, and clinical chemistry, and found that proteomics based models achieved higher average performance for AUROC (Fig. 1E-F), as well as partial AUROC and AUPRC (Fig. S1). Finally, we compare proteomics to an explicit clinical model FINDRISC (*16*), and find that our incident type 2 diabetes prediction model (AUROC = 0.89) outperforms the established FINDRISC diabetes risk model (AUROC = 0.76).

### UKB-trained proteome-disease models generalize well to AoU and FinnGen

To assess model generalizability, we evaluated each UKB-trained model in two external cohorts, with no additional training. For incident disease models, we limited our analysis to prediction of 5-year incidence to account for unequal follow up times across cohorts. As we find that architectures largely show similar performance and generalizability, we highlight ridge regression, as it achieved the highest average performance for both prevalent and incident disease prediction in the majority of cohorts (Table S6). We found similar classification performance between the UKB holdout set and both external cohorts across prevalent and incident disease (Fig. S2; all architectures in Fig. S3-S4). For each disease, we calculated the 95% confidence interval for the change in AUROC between UKB and AoU, and between UKB and FinnGen (Fig. 2).

**Figure 2.**
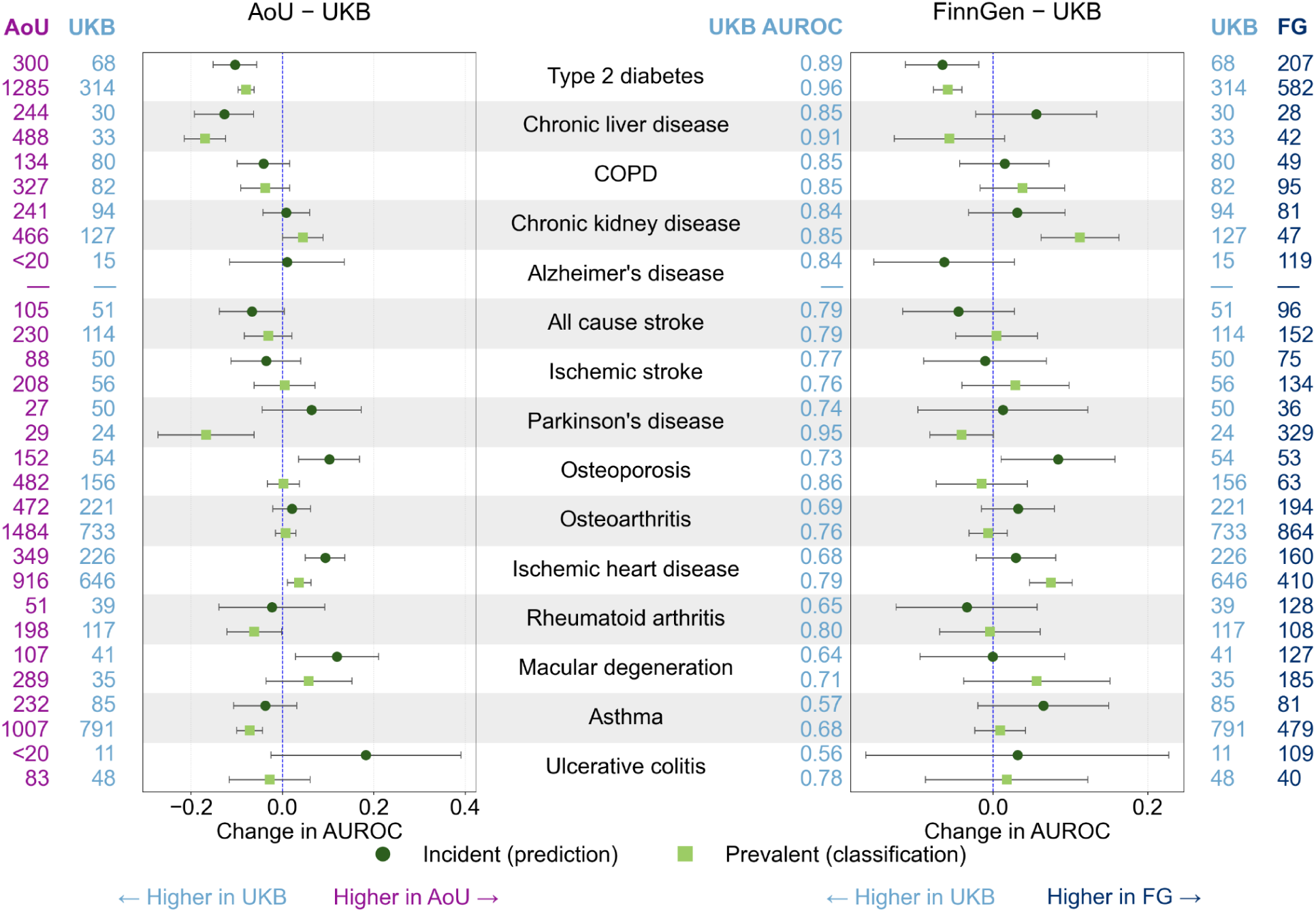
Diseases are predicted accurately and generalize across biobanks. External validation of UKB-trained incident (top row, green circle) and prevalent (bottom row, green square) proteomics disease classification models in FinnGen and AoU. Change in AUROC per disease between UKB and each external test set (AoU left, FinnGen right) is shown for the ridge regression architecture with 95% confidence intervals, ordered by incident UKB performance. Change < 0 indicates better performance in UKB while change > 0 indicates better performance in the external dataset. Case counts for each dataset are shown on the outer columns and UKB holdout AUROC values are in the center.

We defined a model as “generalizing” if the change in AUROC between UKB and the external cohort was greater than or equal to zero, or if its 95% confidence interval included 0. By this definition, all classification models, except type 2 diabetes, generalized in FinnGen. In the AoU cohort, classification model performance was also largely preserved across diseases; however, type 2 diabetes, asthma, chronic liver disease, and Parkinson’s showed a significant decrease in generalizability (Fig. 2). All incident prediction models generalized to FinnGen except type 2 diabetes (Fig. 2), while in AoU, incident prediction models again largely generalized, except type 2 diabetes and chronic liver disease.

### Phenotyping impacts generalizability more than participant demographics and other characteristics

Overall, our results suggest that proteomics based disease models largely generalize across cohorts, with some exceptions. We investigated these exceptions to assess which factors may influence generalizability, including proteomic platform, technical overfitting, age, genetic ancestry, cohort baseline risk factors, environmental risk factors, and phenotyping.

First, Olink proteomic platform and technical overfitting do not appear to substantially degrade model performance. One of the two FinnGen cohorts uses a different panel than UKB, yet all but one disease has consistent performance from UKB to FinnGen across both prevalent and incident disease, making Olink platform and technical overfitting unlikely contributors to model performance. In addition, there are no consistent differences between cohorts when evaluated separately (Fig. S9).

There are several individual diseases that show a lack of generalizability in AoU but not FinnGen, such as prevalent and incident chronic liver disease. Even for type 2 diabetes, which does not generalize in both cohorts, the decreases in performance are larger for AoU than FinnGen. Using a disease definition that requires at least two diagnosis codes in AoU for cases, excluding individuals with only one code, improves AUROC performance for all diseases compared to requiring only one code for both prevalent (t = −7.00, p = 9.2×10^-6^) and incident (t = −6.96, p = 6.6×10^-6^) disease (Fig. 3A-B). For type 2 diabetes, classification AUROC improves by 0.02 and prediction AUROC improves by 0.05. This underscores the impact phenotyping quality can have on model performance. In FinnGen, diabetes case counts are not significantly affected by requiring two diagnosis codes (Supplement).

**Figure 3.**
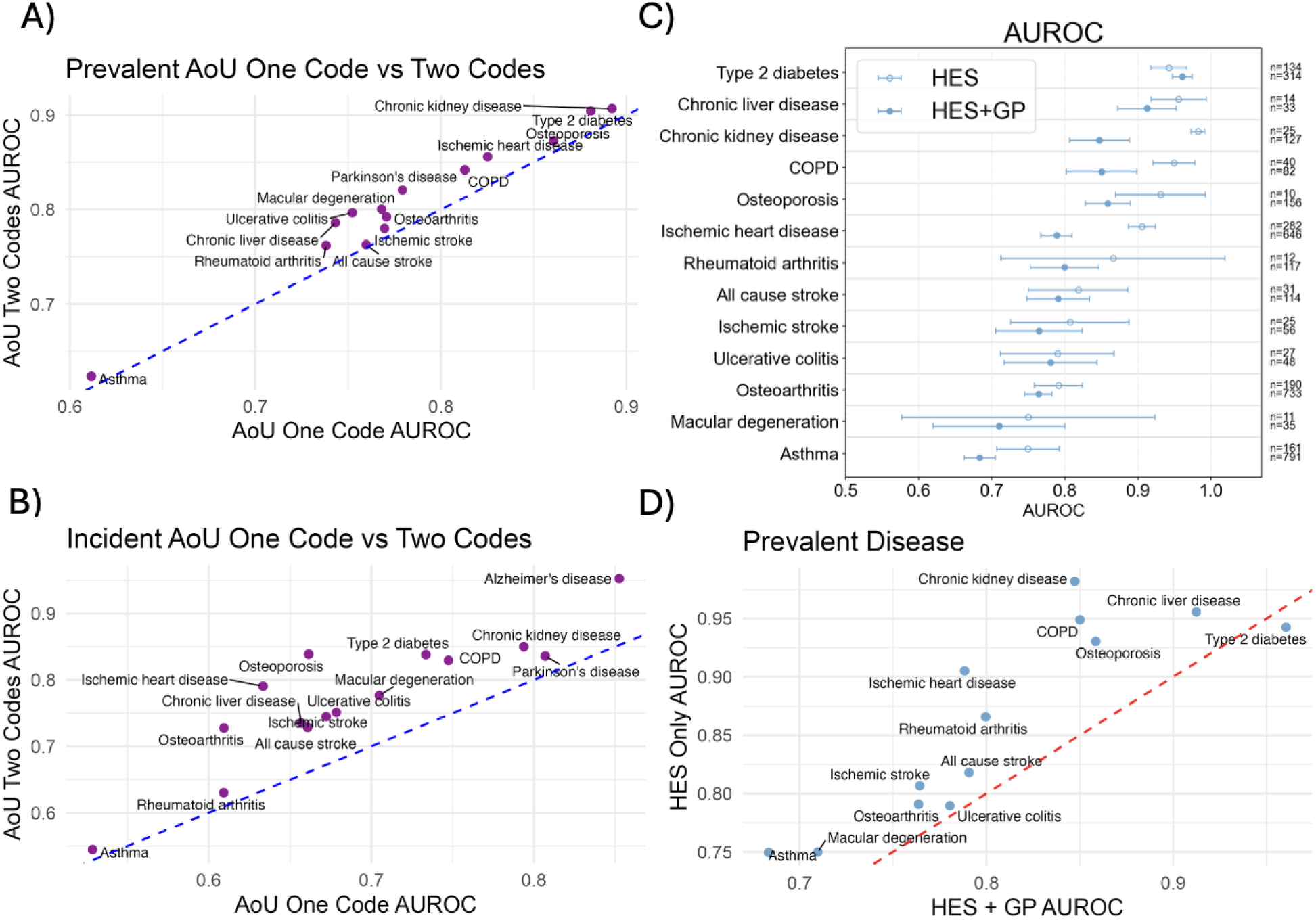
Phenotyping is a major driver of generalizability. We compare prevalent UKB-trained ridge regression performance in AoU when the AoU data is coded by requiring one diagnosis code versus at least two diagnosis codes. For at least two diagnosis codes, individuals with one diagnosis code are excluded from analysis. Model performance is evaluated for AUROC in A) prevalent (t = −7.00, p-value = 9.21 x 10^-6^) and B) 5 year incident (t = −6.96, p-value = 6.61 x 10^-6^). C) We compare prevalent model performance in UKB hold out data when diseases are coded using only hospital admissions (HES) versus coded using a comprehensive set of diagnosis sources including hospital admissions, general practitioner, baseline interview codes, and cause of death (HES+GP). Open dot is HES only, closed dot is HES+GP. Case counts under each data coding scheme are shown. Parkinsonism was excluded due to insufficient case count using HES only (n<5). D) Models built on only hospital admissions data are higher performance than ones with a comprehensive data coding, likely owing to easier prediction of severe outcomes (t = 4.69, p-value = 0.00052).

Furthermore, phenotyping varies between cohorts, although it is largely comparable. Phenotypes are available as ICD9 or ICD10 codes in UKB and AoU; and in various forms (particularly curated endpoints largely based on ICD codes in Risteys) in FinnGen. In addition, UKB differentiates between diagnoses that originated in hospital inpatient admissions versus general practitioner data. Patients who receive a first diagnosis in a hospital setting may have more severe forms of disease that will be easier for a model to learn, leading to higher performance. To test this, we compare the performance of the prevalent classification models in the UKB when we code our UKB hold out test set based only on hospital admissions, versus the comprehensive coding (i.e. including hospital admission, general practitioner, and cause of death) used in original UKB model training. Indeed, when we evaluate the models on hospital admission data only, we see significantly improved average performance across diseases (Fig. 3C). The increase in performance is especially pronounced for prevalent ischemic heart disease and prevalent chronic kidney disease (Fig. 3D). Notably, these are two diseases that show much higher performance in FinnGen compared to other cohorts. Finally, we evaluate the effect of coding type 2 diabetes in UKB using only ICD codes, compared to our current definition that integrates baseline interview codes, insulin use, and HbA1c, and find no significant effect (Supplement)

To further explore the decrease in generalizability for type 2 diabetes classification in AoU compared to other cohorts, we stratify performance by above and below median value for age and BMI, as well as by inferred genetic ancestry for all ancestry groups with at least 1,000 participants for prevalent disease and 500 for incident disease (all shown in Fig. S8), and smoker status (ever vs never) for both classification and prediction models. No significant differences are found between EUR-like, AFR-like, and AMR-like individuals, with small increases in accuracy for SAS-like (Fig 4). However, additional factors such as prevalence, age distribution, and comorbidity prevalence across AoU ancestry subgroups may confound any direct inference. Overall, we find that none of these participant characteristics fully explain the performance decrease (Fig. 4A-B). Furthermore, this trend holds for all disease models (Figs. S6-7).

**Figure 4.**
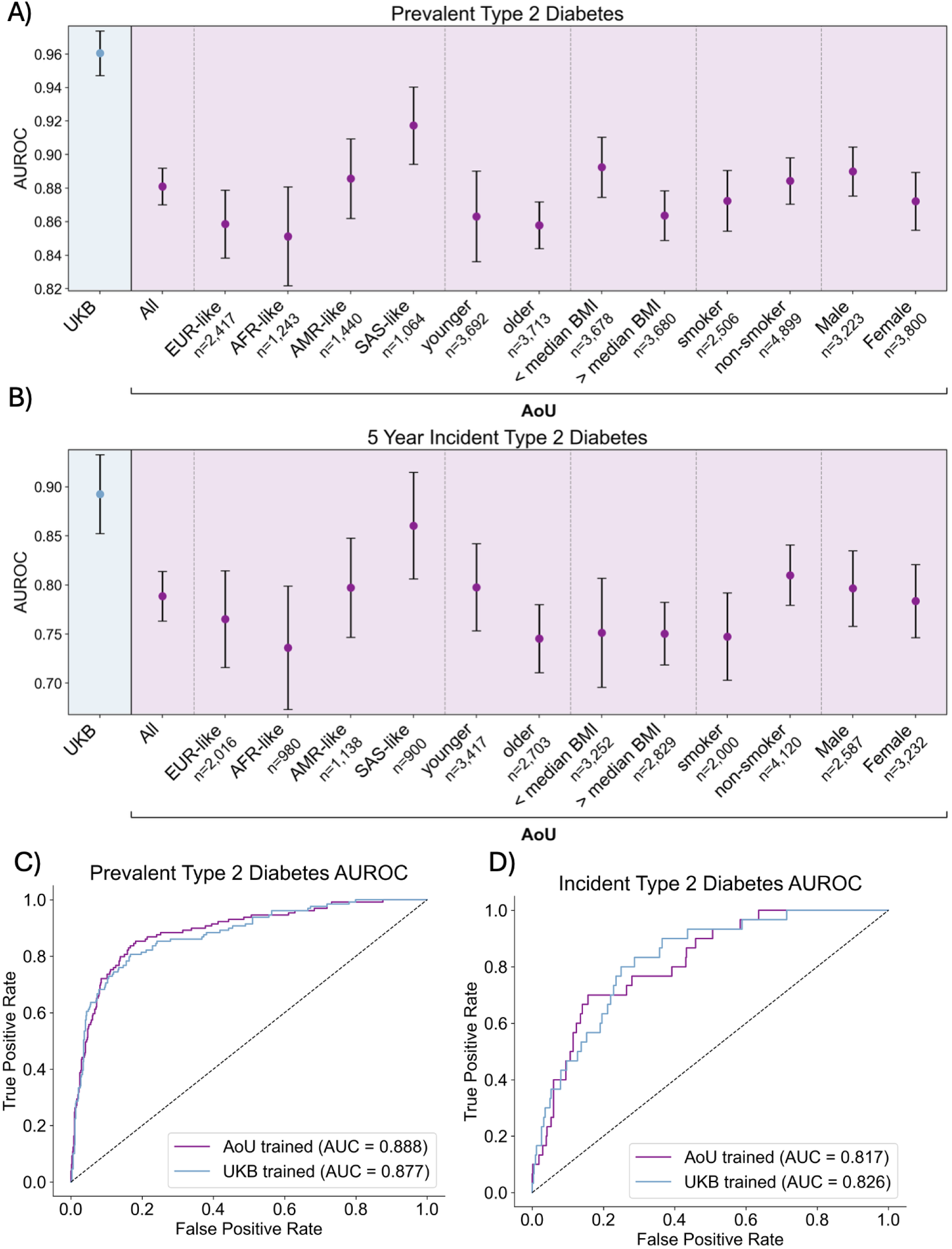
Participant characteristics do not fully explain decreases in generalizability. We evaluate type 2 diabetes model performance stratified by various demographic factors as well as other participant characteristics for A) classification and B) prediction. AoU younger and older are split based on median age (51) for the proteomics cohort. Genetic ancestry groups are derived from AoU definitions via comparison to reference panels. We show data for all ancestry groups with at least 1,000 prevalent participants and 500 incident participants (all groups shown in Fig. S8). AoU smoker is defined as at least 100 lifetime cigarettes. BMI is calculated from AoU participant data, and compared to median BMI of the proteomics cohort (27.4). Individuals missing data for a given strata are excluded from analysis. (C-D): We train a ridge regression model to predict prevalent type 2 diabetes within AoU using a 90%:10% training testing split. We then evaluate performance of the UKB-trained model and the AoU-trained model for AUROC on the 10% AoU hold out set for C) prevalent disease and D) incident disease.

Next, we train prediction and classification type 2 diabetes models on AoU proteomics data, and then evaluate the AoU-trained and UKB-trained models on the same holdout AoU proteomics test set. We find near-identical performance for the AoU-trained and UKB-trained models for classification (Fig. 4C-D) and prediction (Fig. 4E-F), suggesting that the decrease in performance is due to phenotyping in AoU rather than overfitting in UKB. Furthermore, training the AoU model using the full panel of 5k proteins does not improve performance (Fig. S8)

### Specificity of protein models

Because proteomic profiles reflect exposures that can influence many conditions, such as smoking history (*7*), protein → disease associations found via linear regression might be expected to correlate across diseases. Indeed, we find that the univariate effect size vectors are more strongly correlated across pairs of diseases than phenotypic correlation alone can explain (Fig. 5A, Fig. S10A). Thus, we aimed to investigate whether the proteome primarily encodes a generally poor health state or whether these models are specific to a given disease. In a principal components analysis (PCA) of the proteomic data, we find that the first 20 PCs are not predictive of BMI and most participant characteristics (including smoking) with the exception of sex, and they do not maintain the predictive accuracy for any of the diseases studied here (Table S8). The dominant axes of proteomic variation therefore reflect neither disease state nor most exposures, and while we have not yet ruled out any technical confounders (*17*), these results do not identify a single general health component as the primary driver of proteomic profiles; instead, disease- and exposure-related signals may reside in more specific proteins.

**Figure 5.**
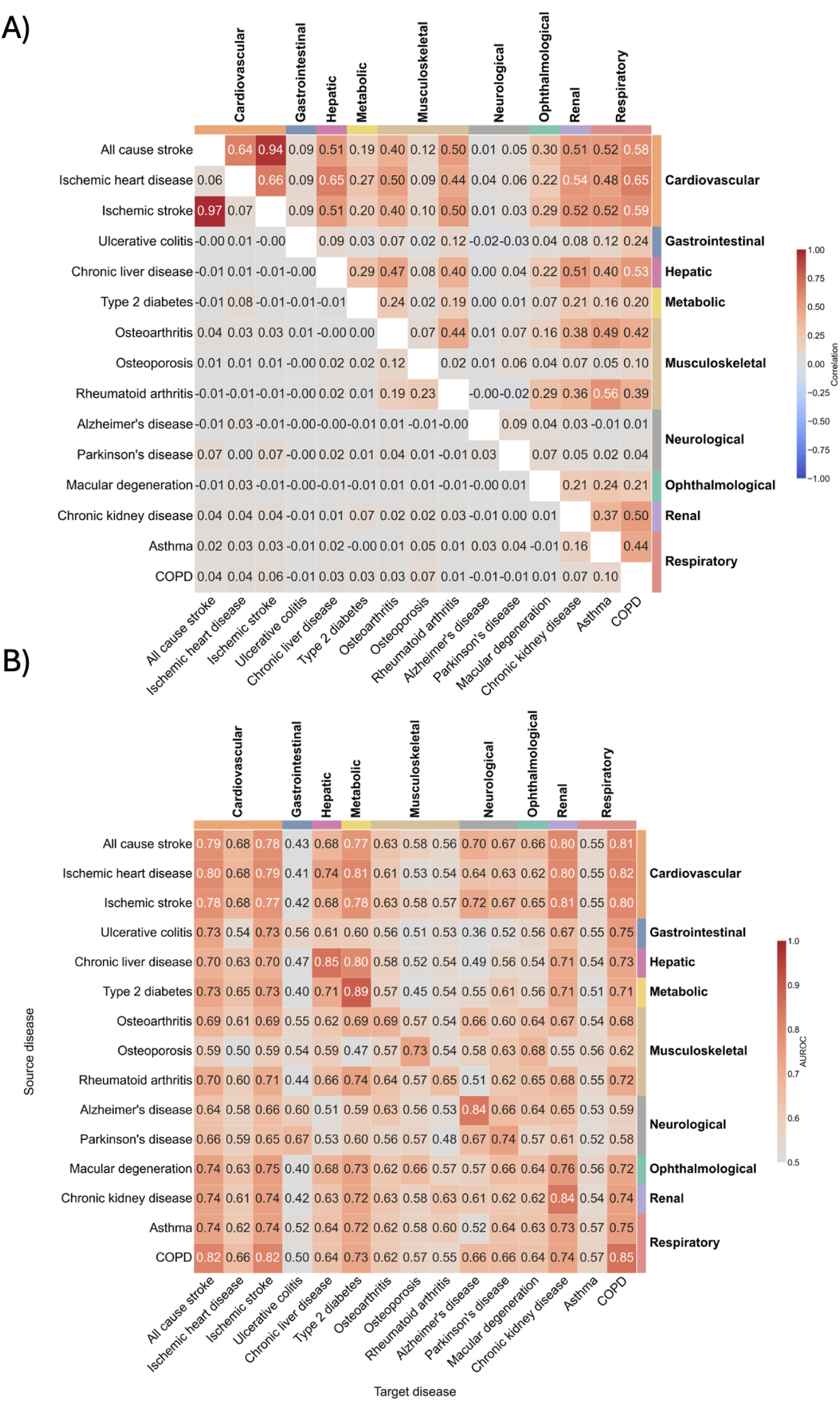
Incident disease models show similar signatures but are disease-specific. A) Combined correlation heatmaps for incident diseases. Pearson correlations between binary disease indicators (phenotypic correlation, lower triangle) and disease-specific coefficient vectors derived from univariate proteomic association analyses (β-vector correlation, upper triangle). For each disease, coefficient vectors span 2,920 plasma proteins from the UK Biobank Olink panel; coefficients with an adjusted p-value ≥ 0.01 were set to zero prior to correlation. B) Performance of L2 ridge regression models applied to all other diseases and evaluated using AUROC for incident diseases. Source disease is the disease used to evaluate and target disease is the disease being predicted by the source model.

Having ruled out a single dominant health axis, we next asked how much signal is shared across disease-specific models. Using each model to predict every other disease, we find that models generally retain some predictive performance across a range of diseases, implying overlap in the proteomic signature for different diseases (Fig. 5B, Fig. S10B). In most cases, however, each disease is best predicted by the model on which it was trained, with the exception of some instances where the target disease has a smaller sample size. A PheWAS between each protein score and all available phenotypes further shows that the top associations are domain-specific (Fig. S11): for example, HbA1c is in the top 10 associations for both prevalent and incident type 2 diabetes. Together, these results suggest that although there is overlap among disease proteomic signatures, each model retains domain-specific tuning rather than reducing to a common readout of poor health.

Finally, to investigate whether the protein disease scores captured heritable components of disease liability, we computed their genome-wide association profiles as molecular-trait proxies and assessed heritability and genetic correlations using LD score regression (*18*). These analyses showed modest and consistent SNP heritability (h^2^ range = 0.047-0.139), indicating that the predicted protein scores captured a reproducible polygenic component rather than only environmental or measurement variation. We estimated genetic correlations with 30 UK Biobank trait GWAS spanning ten broad phenotype categories, and across 870 pairwise genetic correlation tests, 448 associations (51%) remained significant after Bonferroni correction (p < 5.7 x 10^-5^), with several biologically coherent patterns (Fig. S12). For instance, cardiovascular disease scores overlapped most strongly with cardiovascular traits (mean r_g_ = 0.46; 94% Bonferroni-significant), and renal disease scores showed strong genetic overlap with kidney-function biomarkers such as chronic kidney disease, eGFR, and creatinine (mean |r_g_| = 0.73; 100% Bonferroni-significant), consistent with disease-specific signal captured by each protein score. Overall, genetic correlations are stronger than raw phenotypic correlations but the two modalities have concordant directionality.

## Discussion

We built classification and prediction models for 15 chronic diseases that are major causes of morbidity and mortality, using proteomics data from UKB, which achieve high performance in two external cohorts of proteomics data, AoU and FinnGen. To our knowledge, this is the largest cross-biobank evaluation of the generalizability of proteomics-based disease prediction models, particularly across diverse environments in the United States. This complements prior large-scale cross-cohort work on protein-level associations (e.g., pQTL studies (*19*)) by focusing specifically on model performance for disease classification and prediction across multiple cohorts. Overall, model generalizability was high across the majority of diseases and cohorts; we investigate the exceptions and factors that may drive them. In particular, performance for type 2 diabetes models decreased in external cohorts, particularly AoU. We find that participant characteristics such as age, sex, genetic ancestry, BMI, and smoking status are not primarily responsible for decreases in disease risk model performance, a notable difference from other data modalities such as polygenic risk scores. However, we note that our subgroup comparisons may be underpowered to detect moderate effect sizes. We therefore cannot fully rule out that such participant characteristics contribute to cross-cohort performance differences, even though they do not appear to fully explain them.

Instead, we find that phenotype definition is an important contribution to model performance, as requiring at least two diagnosis codes significantly improves performance for all diseases in AoU. This implies phenotyping quality in AoU is a major driver of model performance in this cohort. Consistent with this, models evaluated on hospital-admission diagnoses alone in UKB outperformed those using comprehensive coding, reflecting the easier prediction of more severe disease captured in inpatient records. While requiring at least two diagnosis codes reduces phenotype noise, it is not without its limitations, such as excluding some true cases and enriching for more severe cases. Future work should consider evaluating differences in phenotyping quality and depth between biobanks and clinically curated datasets. Moreover, the number of diagnosis codes a participant accumulates also reflects the frequency and continuity of their healthcare access and utilization rather than disease presence or severity alone. Because healthcare access and utilization can vary across cohorts, this phenotyping approach may differentially affect case ascertainment across subgroups, potentially undercounting true cases among participants with less frequent healthcare contact. Future work should evaluate whether performance gains from stricter diagnosis-code thresholds hold consistently across subgroups.

A related limitation is that we do not exhaustively assess which variables may lead to differences in performance across cohorts, as variables such as household income, insurance status, and other correlates of healthcare access are not necessarily collected or comparable in the same way across the national health systems represented by UKB, AoU, and FinnGen. In a forthcoming companion manuscript (Jee et al., 2026, in preparation), we assess 108 of these metrics for proteomics scores of exposures. These differences could themselves contribute to the discrepancies we observe here, particularly for a disease like type 2 diabetes where diagnosis and severity are closely related to healthcare contact and overall health (e.g. BMI).

Future work should evaluate the relative contributions of healthcare contact versus general health characteristics such as BMI and smoking status in driving disease prediction. Overall we find that most diseases generalize, with a focus on type 2 diabetes as an exception. This study cannot fully distinguish biological non-generalizability from social and healthcare system-level causes. Identifying which specific structural or data-collection differences may limit generalizability remains an important direction for future work.

Simple architectures (ridge regression) performed at least as well as more complex architectures across nearly all disease-cohort combinations (Table S6, S7) for both internal performance or cross-cohort generalizability. Beyond portability, linear models with a single weight per protein carry substantially less risk of encoding individual-level training data than high-capacity alternatives, and are therefore easier to share and deploy across biobanks without transferring individual-level data. Future work that extends these using Mendelian Randomization or mediation analysis will enable more interpretable models that better characterize the biology of these diseases.

Our results show good generalizability despite the application to two Olink platforms across cohorts with independent normalization, implying that this technical element does not have a large effect on model performance. However, we notably do not compare Olink to other proteomics platforms, which may influence generalizability. Because Olink and affinity- or mass-spectrometry-based platforms differ in both protein coverage and measurement scale, models trained on one may require recalibration rather than transferring directly. Cross-platform validation is therefore an important next step for establishing whether proteomic models are portable across assay technologies as well as across cohorts.

Biobanks differ substantially in phenotyping quality and sources of disease data. Our results indicate that this heterogeneity, rather than the proteomic assay or cohort composition, is the principal barrier to cross-cohort generalizability. A more nuanced investigation of disease severity and outcome definitions may lead to more accurate and interpretable models, particularly as longitudinal data become available. In addition, the genome-wide genetic correlations between protein disease scores and sociodemographic and behavioral traits were derived solely from UKB and have not been evaluated in AoU or FinnGen. Given substantial differences among these biobanks in participant demographics and recruitment, as well as in the methods used to collect sociodemographic and behavioral data, these genetic correlations should not be assumed to generalize across biobanks. More broadly, our findings indicate that the reproducibility of proteomic prediction depends as much if not more on the infrastructure of phenotype ascertainment as on human biology.

## Supporting information

supplementary results

## Data Availability

This study used data from the All of Us Research Program Controlled Tier Dataset version 8, available to authorized users on the Researcher Workbench. UK Biobank data are available through a procedure described at https://www.ukbiobank.ac.uk/enable-your-research. FinnGen data can be accessed through Fingenious services (https://site.fingenious.fi/en/) managed by FINBB. Finnish Health Register data can be applied for from Findata (https://findata.fi/en/data/). Code developed for this project in AoU is available as a featured workspace (https://workbench.verily.com/workspaces/proteomics).

## Acknowledgements

We gratefully acknowledge participants in the UK Biobank, All of Us, and FinnGen for their contributions, without whom this research would not have been possible. We also thank the National Institutes of Health’s All of Us Research Program for making available the participant data, samples, and cohorts examined in this study. We also thank the leadership of the UK Biobank and FinnGen. K.J.K. was supported by grants from the National Institutes of Health, including the National Human Genome Research Institute under Award Number UG3HG014379, and the Broad Next Generation Fund. K.J.K. and B.M.N. were supported by Novo Nordisk Foundation (NNF21SA0072102). M.A.A. was supported by funds received from the Firuza Foundation. Multiomics data generation in the All of Us Research Program was supported by the National Institutes of Health under OT2OD038121.

## Data and Code Availability

This study used data from the All of Us Research Program’s Controlled Tier Dataset version 8, available to authorized users on the Researcher Workbench. UK Biobank data are available through a procedure described at https://www.ukbiobank.ac.uk/enable-your-research. FinnGen data can be accessed through Fingenious services (https://site.fingenious.fi/en/) managed by FINBB. Finnish Health Register data can be applied for from Findata (https://findata.fi/en/data/). Code developed for this project in AoU is available as a featured workspace (https://workbench.verily.com/workspaces/proteomics).

## Notes

### Competing Interest Statement

Mitja I. Kurki is co-founder and paid scientific advisor of Switchpoint Bio
Benjamin M. Neale is a member of the scientific advisory board at Deep Genomics, Vesalius Therapeutics, Camp4 Therapeutics and Aluco BioSciences Inc.
Michael E. Talkowski receives research funding and support from the NIH, SFARI, the Gates Foundation, Massachusetts General Hospital, Philanthropic initiatives, and the Foundation for Prader-Willi Research and industry funding and/or reagents from Ionis Pharmaceuticals, Illumina, Pacific Biosciences, and Microsoft. He is also a consultant and computational SAB member for BridgeBio Inc. and co-founder of First Genomic Insights, LLC.
Stephen B. Montgomery is an advisor to BridgeBio, MyOme and PhiTech
Mark J. Daly has a financial interest in Maze Therapeutics, a company Focusing on genetically informed therapies for common diseases such as chronic kidney disease. The interests of Dr. Daly were reviewed and are managed by MGH and Mass General Brigham in accordance with their conflict-of interest policies.
Konrad J. Karczewski is a member of the scientific advisory board of Nurture Genomics

### Author Declarations

Adults 18 years and older who have the capacity to consent and reside in the USA or a US territory at present are eligible to participate in the All of Us Research Program. Informed consent for all participants is conducted in person or through an eConsent platform that includes primary consent, HIPAA Authorization for Research use of EHRs and other external health data, and Consent for Return of Genomic Results. This secondary data analysis research did not require IRB approval because it does not involve human participants. All data is now available on the All of Us Researcher Workbench. FinnGen use of national healthcare data and biospecimens is in compliance with national (Finland's Act on the Secondary Use of Health and Social Data) and European (the General Data Protection Regulation) regulations. These regulations prohibit the distribution of individual-level data. This study used data from the UK Biobank. UK Biobank has approval from the North West Multi-centre Research Ethics Committee (MREC) as a Research Tissue Bank (RTB) approval. This approval means that researchers do not require separate ethical clearance and can operate under the RTB approval.

