## supplementary results for "Generalizability of proteomic risk prediction across biobanks reveals dependence on phenotype definitions"

### Supplementary information for “Generalizability of proteomic risk prediction across biobanks reveals dependence on phenotype definitions”

**Proteomics cohorts** 3
**Outcomes** 3
**Biobank data processing** 4
UKB proteomics 4
UKB clinical data 5
FinnGen proteomics 5
AoU proteomics 6
Handling missing proteins across cohorts 6
**Model training** 6
Logistic regression 7
LightGBM (LGBM) 7
Multilayer perceptron (MLP) 7
Transformer model pretraining 7
Transformer disease-specific fine-tuning 8
HES vs HES+GP Analysis 8
**Model evaluation** 8
Clinical data versus proteomics 8
Figure S1. Proteomics vs clinical data 10
FINDRISC model comparison 11
Model evaluation across cohorts 11
Figure S2. AUROC external cohorts vs UKB 11
Figure S3. Prevalent classification model performance for all architectures 14
Figure S4. Incident prediction model performance for all architectures 16
Figure S5. Partial AUROC external cohorts 17
**Factors influencing generalizability** 17
Figure S6. AoU classification performance stratified by participant characteristics 19
Figure S7. AoU prediction performance stratified by participant characteristics 20
Figure S8. AoU prediction performance stratified by genetic ancestry 21
Figure S9. AoU type 2 diabetes prediction 23
Figure S10. FinnGen subcohort performance 25
**UKB cohort structure** 25
Figure S11. PCA of UKB proteomics 27
Figure S12. Random forest from UKB principal components 29
**Proteomic univariate analysis** 29
**Model interpretation** 29
Figure S13. Prevalent model specificity 31
Figure S14. PheWAS of incident type 2 diabetes 31
Genome-wide association analyses of protein scores 32

Genetic correlation analysis 32
Figure S15. Phenotype associations vs GWAS signal 34
**Effect of training case count on model performance** **35**
Figure S16. Effect of training case count on type 2 diabetes model performance 35
**Supplementary Tables** **36**
Table S1. Curated disease phenotypes in UKB and AoU 36
Table S2. FinnGen phenotype mapping 37
Table S3. LGBM hyperparameter search space 39
Table S4. MLP hyperparameter search space 39
Table S5. FINDRISC criteria mapped to UKB fields 40
Table S6. Average model performance across architectures 41
Table S7. Comparison of all architectures to ridge regression 41
Table S8. Predictive power of top 20 PCs vs proteomics data 42
References 43

#### Proteomics cohorts

We evaluated three large population-based cohorts: UK Biobank (UKB), All of Us (AoU), and FinnGen. UKB is a prospective cohort of approximately 500,000 participants recruited in the United Kingdom between 2006–2010, with extensive phenotyping, genotyping, and linked health records. Proteomics data is available using the Olink Explore 3072 platform for a randomly selected subset of 46,595 UKB participants at the baseline visit, 6,376 individuals selected by the UKB-PPP consortia, and 1,268 individuals from a COVID-19 study(*1*).

The AoU Research Program is a diverse cohort targeting over 1 million participants in the United States, with strong representation of historically underrepresented populations and integration of electronic health records, survey data, and biospecimens. Olink Explore HT proteomics data is available for a subset of individuals as part of the multiomics demo pilot, selected to maximize ancestral diversity (Padhi et al. 2026, in preparation).

Finally, FinnGen combines genetic data from Finnish biobanks with nationwide health registry data, currently including over 500,000 individuals. FinnGen contains two main cohorts of proteomics data. The first cohort (Cohort A, n=1,925) uses the Olink Explore 3072 panel and includes primarily healthy blood donors who carry rare genetic variants. The second cohort (Cohort B, n=3,940) uses the Olink Explore HT platform and measures the proteome for 1,608 healthy blood donors as well as additional individuals from disease specific task groups.

#### Outcomes

Diseases were coded as prevalent if the first disease event date was prior to the proteomics sample collection date and as incident if the first occurrence was after sample collection. Prevalent cases were excluded as cases or controls from incident prediction tasks. Diseases with fewer than 100 total cases in the UKB were excluded. In each external cohort, disease case counts are reported in compliance with biobank specific policies (<=5 in FinnGen, <=20 in AoU). Diseases were excluded from analysis for external cohorts if there were fewer than two cases.

Disease diagnoses in the UK Biobank were ascertained using ICD9 and ICD10 diagnosis codes and diagnosis dates from linked hospital inpatient, primary care, cancer registry, and death registry records with the addition of baseline interview codes and measurements for some diseases. The full set of criteria used for each disease can be found in Table S1. Primary care read codes were mapped to corresponding ICD codes using the UK Biobank-provided lookup table.

Disease diagnoses in AoU were made using the same ICD9 and ICD10 code combinations as in the UKB (Table S1). ICD codes are available in All of Us for individuals who opt to link their electronic health record. We exclude individuals with no linked record (n=2,602).

Disease diagnoses in FinnGen are available from a broad range of national registries. In particular, hospital inpatient and outpatient diagnoses are available from the Finnish Social and Healthcare Notification System (HILMO) dating back to 1969. This databank is maintained for accuracy by the Finnish Institute for Health and Welfare. FinnGen has released a range of disease endpoints that integrates ICD8, ICD9, and ICD10 codes from hospital discharges and cause of death registries along with medication and drug reimbursement information to define a comprehensive case definition. We map our UKB disease definitions to FinnGen endpoints using ICD9 and ICD10 codes. The complete set of endpoints along with corresponding ICD codes for each disease is shown in Table S2.

#### Biobank data processing

In all cohorts, the released proteomics data are based on the Olink NPX unit on a log2 scale. Within each cohort, protein expression is normalized by minmax scaling followed by median centering, per protein. The Olink HT panel is nearly a superset of the Explore 3072 panel, in which up to 2,844 of the 2,920 proteins of the Explore 3072 panel are found in the HT panel.

##### UKB proteomics

The full UKB proteomics cohort measured 2,923 proteins using the Olink Explore 3072 platform (*1*). We exclude 3 proteins (GLIPR1, NPM1, PCOLCE) with >20% missingness for a total of 2,920 unique proteins. Since the transformer architecture can inherently handle missing values, we do not impute the data for transformer training. For all other architectures, the proteomics data is imputed using the miceforest Python package as previously described (*2*).

UKB clinical data

Complete blood counts and clinical chemistry were run on the same blood sample as the proteomics in the UKB. We remove highly correlated blood count metrics and include 14 metrics in our final analysis, as described in past work (*3*). The metrics are leukocyte count, haematocrit percentage, platelet count, monocyte percentage, neutrophil percentage, eosinophil percentage, basophil percentage, reticulocyte percentage, high light scatter reticulocyte percentage, immature reticulocyte fraction, mean corpuscular erythrocyte volume, mean reticulocyte volume, mean sphered cell volume, and mean platelet volume. For clinical chemistry, we use 30 metrics: Alanine aminotransferase, Albumin, Alkaline phosphatase, Apolipoprotein A, Apolipoprotein B, Aspartate aminotransferase, C-reactive protein, Calcium, Cholesterol, Creatinine, Cystatin C, Direct bilirubin, Gamma glutamyltransferase, Glucose, Glycated haemoglobin (HbA1c), HDL cholesterol, IGF-1, LDL direct, Lipoprotein A, Oestradiol, Phosphate, Rheumatoid factor, Sex hormone-binding globulin (SHBG), Testosterone, Total bilirubin, Total protein, Triglycerides, Urate, Urea, and Vitamin D.

Missing values for blood counts and clinical chemistry are imputed using mean value imputation. The final clinical dataset includes age, sex, 14 blood count metrics, and 30 biochemistry metrics. Each metric, except for sex, is normalized using the same method as for the proteomics data, minmax scaling followed by median centering.

FinnGen proteomics

The FinnGen cohort A uses the Olink Explore 3072 to measure plasma proteomics in 1990 individuals, primarily healthy blood donors. Data imputation was done using miceforest, as described for UKB. We use the set of 1925 individuals with non-missing plasma collection dates and health information in FinnGen longitudinal endpoint files.

The FinnGen cohort B used the Olink Explore HT panel to measure ~5,300 proteins for 1,608 healthy blood donors along with individuals from disease specific task forces. It includes kidney disease (n=945), IBD (n=268), rheumatic diseases (n=276), metabolic disease (n=133), Parkinson's (n=335), pulmonary diseases (n=46), eye diseases (n=162), rare variant carriers (n=500), chromosomal abnormalities (n=153), gender identity disorders (n=61), and younger individuals (n=53). The final version of the data released after QC measures 5,420 proteins.

We use only the subset of proteins included in the UKB panel. Proteins were mapped across cohorts using the UniProt accession number. Protein expression was normalized using minmax scaling followed by median centering for each cohort separately. Data from both FinnGen cohorts were combined for final model evaluation.  Sub analyses were also run on the two cohorts separately (Fig S10).

AoU proteomics

The AoU cohort contains a set of individuals selected to maximize ancestral diversity and include populations traditionally underrepresented in biomedical research (Padhi et al. 2026, in preparation). Samples were selected for proteomic analysis from the AoU long-read cohort. This cohort includes self-identified African American or Black participants as well as individuals of self-identified American Indian or Alaska Native (AI/AN) heritage. Additional samples were selected for ancestry representation across six computed continental ancestry groups (AFR, AMR, EAS, EUR, MID, SAS). A total of 29 individuals were missing biosample collection dates for the proteomics data and were excluded. Individuals with no EHR were excluded. Proteins were mapped from the provided Olink id to UniProt ID, which was then used to align with the UKB proteomics data.

Handling missing proteins across cohorts

Some proteins present in the UKB panel are not present in the other cohorts: in the FinnGen cohort A, there are 21 such proteins, in the FinnGen cohort B there are 76 such proteins, and in All of Us there are 80 such proteins. The expression of these proteins was input as missing for the transformer architecture, which inherently imputes missing data, and were imputed with a normalized value of 0 for all other architectures, which corresponds to median value imputation under the minmax scaling followed by median centering normalization process.

### Model training

We created a single, random 90% train (n=47,723) 10% test (n=5,303) split of the UKB proteomics data that is used across all architecture types. In all cases, final performance was evaluated on the held-out test set and external cohorts.

Logistic regression

We trained separate L1- and L2-regularized models using scikit-learn's (version 1.7.1) LogisticRegression module with 5 fold cross validation, the saga solver, and a maximum of 1000 iterations to convergence. The regularization strength was tuned using grid search for parameter space [0.001, 0.005, 0.01, 0.05, 0.1, 0.5, 1, 2, 5, 10, 50, 100].

LightGBM (LGBM)

Gradient-boosted decision trees were tuned with Optuna over 50 trials, optimizing mean AUROC via 5-fold stratified cross-validation on the training set over a wide range of hyperparameter values. The search space included learning rate, number of estimators, number of leaves, max depth, min data in leaf, feature fraction, bagging fraction/frequency, L1/L2 penalties, and min_gain_to_split (Table S3). We also apply class weighting using scale_pos_weight, setting the value to the ratio of controls to cases. After hyperparameter selection, the final model was trained on a stratified 90% subset of the training data, using the remaining 10% as an internal validation set for early stopping.

Multilayer perceptron (MLP)

We tuned a Keras Sequential MLP with Optuna for 50 trials using 5-fold stratified CV on the training set. The search (Table S4) varied the number of hidden layers (3–6), units per layer (32–1024, log-scaled), activation (relu/selu/elu), dropout (0.10–0.50), learning rate ($10^{-4}$–$10^{-2}$, Adam optimizer), and batch size (64/128/256/512). Each CV fold trained up to 50 epochs with early stopping on validation AUROC (patience 10). The selected configuration was retrained on the full training set with 10% internal validation for early stopping.

Transformer model pretraining

The single training set described above (90% of the original data), was further split so that 80% of the original data was used for training and 10% was used for validation for the transformer. We pretrained a transformer on the tabular proteomics data using a masked-value reconstruction objective. For each individual, 15% of observed (non-missing) protein measurements were randomly masked per batch; missing values were also treated as masked. Masked inputs were replaced by a constant mask token (0.0), and a Boolean mask indicator was supplied to the model for each position. The network predicts the original normalized protein expression value for every of the 2,920 proteins, with mean squared error as the loss function, computed only over the artificially masked positions.

The transformer encoder has 6 layers, 2 attention heads per layer, model dimension ($d_{model}$) 512, dropout 0.1, and feed-forward dimension 4*$d_{model}$. Each scalar feature value is linearly embedded to $d_{model}$ dimension vector and then summed with a learned feature embedding and a 2-state mask embedding. Finally, the encoder output is projected back to one scalar value per feature. We trained with AdamW (learning rate $10^{-4}$, default betas and weight decay), gradient-clipping at 1.0, batch size 128, and a 50-epoch schedule: 10 epochs of linear warm-up to $10^{-4}$ followed by cosine decay for the remaining 40 epochs. The checkpoint with the lowest validation loss was retained.

Transformer disease-specific fine-tuning

We fine tune separate models for each of the diseases using the pretrained transformer by adding a new classification head. We mean-pool the final hidden states across features and pass the pooled vector through the classification head to obtain a logit. No weights are frozen during fine tuning so the entire encoder is tuned. Models were fine tuned for 10 epochs with AdamW (learning rate $10^{-5}$), batch size 128, and binary cross-entropy with logit loss. The held-out test split (original 10% described above) was reserved for final evaluation and was not used in training or model selection.

HES vs HES+GP Analysis

We compare prevalent model performance on our UKB hold out set when the hold out set is coded considering only hospital admissions data, versus hospital admissions and general practitioner data. We exclude diseases without at least 5 cases in the hospital admissions only based coding.

### Model evaluation

Clinical data versus proteomics

As a comparator with the proteomics based models, we also trained prevalent and 5 year incident disease prediction models based on the clinical data (age, sex, complete blood cell count, and clinical chemistry) described earlier, using an identical train/test split as the proteomics models. The model training process was the same as described for proteomics based prediction for L1 and L2 penalized logistic regressions, LGBMs, and MLPs. We did not train a transformer on clinical data due to the low number of features in the clinical dataset. We compare model performance using AUROC, partial AUROC, and AUPRC, highlighting the ridge regression models (Fig. S1). For prevalent disease, we find that proteomics significantly outperforms clinical data in a pairwise t test by disease for all three metrics (AUROC p = 0.007514, partial AUROC p = 0.007861, AUPRC p = 0.01273). For incident disease, proteomics outperforms clinical data on average, but does not reach significance. In all metrics, and for both prevalent and incident disease, the clinical data models outperform the proteomic models for type 2 diabetes. This is likely due to the inclusion of HbA1c, a diagnostic biomarker for type 2 diabetes, as well as glucose in the clinical chemistry data. Chronic kidney and chronic liver disease also show higher performance in AURPC with clinical data. These diseases also have significant biomarkers represented in clinical chemistry sets due to the inclusion of creatinine, alanine aminotransferase, and aspartate aminotransferase.

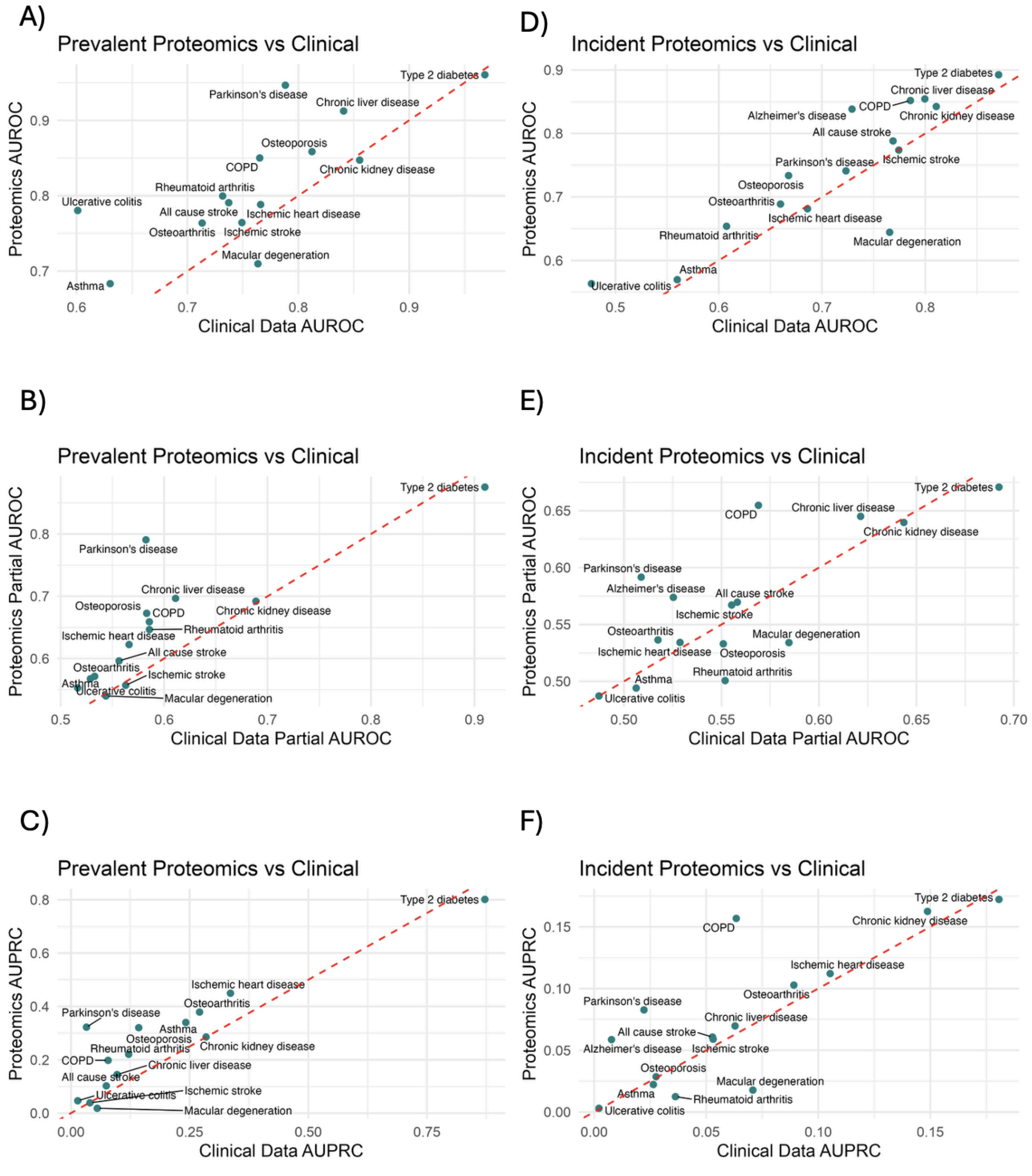


**Figure S1. Proteomics vs clinical data**

Using the same training testing split as proteomics models, we train models using baseline clinical data using ridge regression. For prevalent disease, we compare proteomics and clinical data performance in hold out test data for A) AUROC (t = 3.16, p = 0.0075), B) partial AUROC with maximum 5% false positive rate (t = 3.13, p = 0.0078), and C) AUPRC (t = 2.88, p = 0.012). We also train 5 year incident prediction models and compare models for D) AUROC (t = 2.12, p = 0.052), E) partial AUROC (t = 0.84, p = 0.41) and F) AUPRC (t = 1.27, p = 0.22).

FINDRISC model comparison

We compare our proteomics based incident 5 year type 2 diabetes prediction model with the FINDRISC model. We translate UKB fields to the relevant FINDRISC questions and then add all points per patient. Table S5 shows the conversion used between FINDRISC question, UKB fields, and risk score points.

Model evaluation across cohorts

Models were evaluated with AUROC (Fig S2), partial AUROC with 5% false positive rate, and AUPRC with 95% confidence intervals for each metric on hold out test data. Confidence intervals (CIs) for the AUROC were obtained using the confidenceinterval package (version 1.0.5) which implements the DeLong analytical method to calculate AUROC confidence intervals. Confidence intervals for the difference in performance between UKB and each external validation set were obtained from the individual cohort confidence intervals. The standard error for each cohort was estimated as CI width/(1.96*2). The standard error for the difference ($se_{diff}$) was then calculated as sqrt($se_{ukb}^2$ + $se_{external}^2$). Finally, the confidence interval was calculated as +/- 1.96*$se_{diff}$ around the point estimate of the difference.

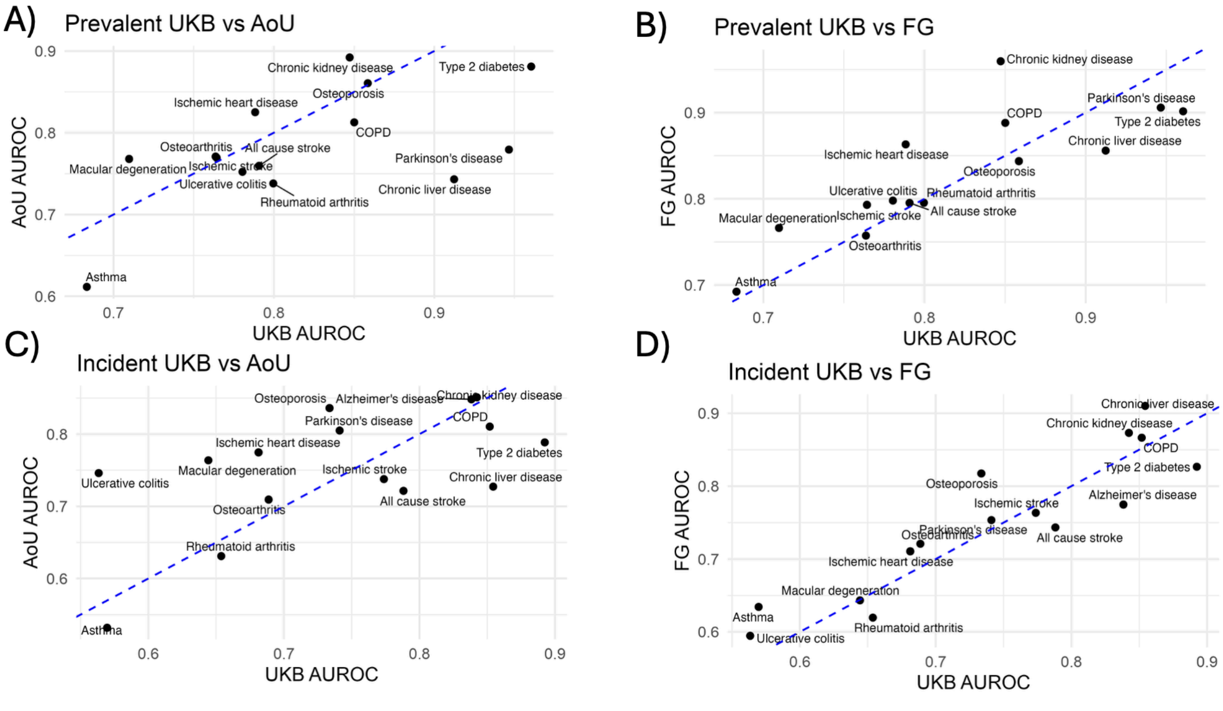


**Figure S2. AUROC external cohorts vs UKB**

We evaluate the AUROC for ridge regression classification models in A) UKB vs AoU and B) UKB vs FinnGen and for prediction models in C) UKB vs AoU and D) UKB vs FinnGen

For proteomics modeling, we evaluated five different architectures, as described above. All models are trained in the UKB, and within the UKB hold out set, we see no significant differences between architecture AUROC performance for both prevalent and incident disease. This trend largely persists in external validation cohorts, FinnGen and All of Us, for both prevalent and incident disease, with a few exceptions (Fig. S3, Fig S4, Table S6, Table S7).

Parkinson's classification is notable for high variability in generalizability between the different architectures in AoU (AUROC range 0.56-0.78). In UKB hold out, both logistic regression models perform better than the LGBM and MLP architectures, but in AoU they perform similarly to the other architectures and therefore do not generalize their increased performance. The logistic regression models and transformer outperform other architectures for prevalent Parkinson's and macular degeneration in FinnGen. The lasso logistic regression and transformer outperform other architectures for incident Alzheimer's in FinnGen.

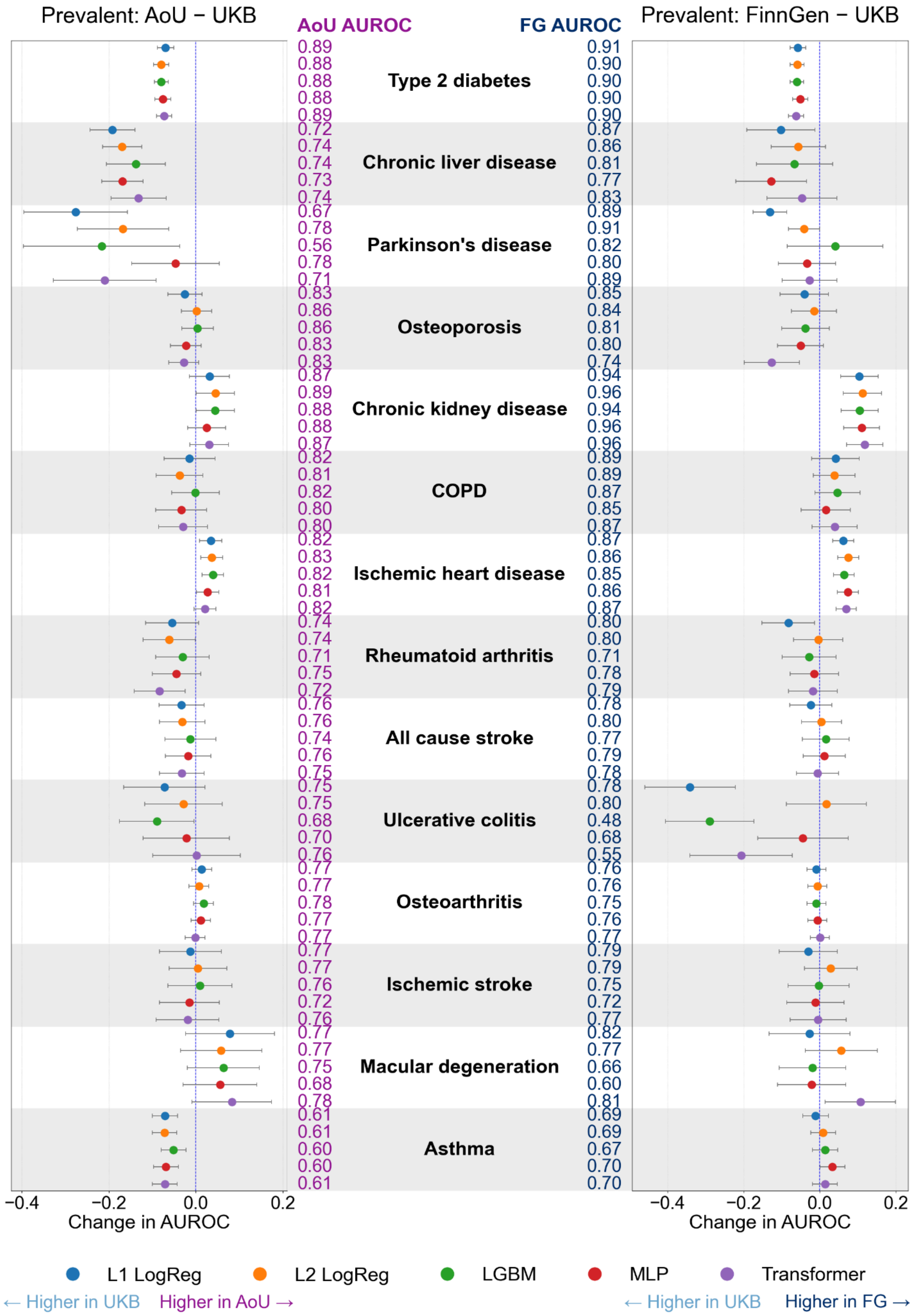

Prevalent: AoU − UKB
AoU AUROC
FG AUROC
Prevalent: FinnGen − UKB
Type 2 diabetes
0.89 0.88 0.88 0.88 0.89
0.91 0.90 0.90 0.90 0.90
Chronic liver disease
0.72 0.74 0.74 0.73 0.74
0.87 0.86 0.81 0.77 0.83
Parkinson's disease
0.67 0.78 0.56 0.78 0.71
0.89 0.91 0.82 0.80 0.89
Osteoporosis
0.83 0.86 0.86 0.83 0.83
0.85 0.84 0.81 0.80 0.74
Chronic kidney disease
0.87 0.89 0.88 0.88 0.87
0.94 0.96 0.94 0.96 0.96
COPD
0.82 0.81 0.82 0.80 0.80
0.89 0.89 0.87 0.85 0.87
Ischemic heart disease
0.82 0.83 0.82 0.81 0.82
0.87 0.86 0.85 0.86 0.87
Rheumatoid arthritis
0.74 0.74 0.71 0.75 0.72
0.80 0.80 0.71 0.78 0.79
All cause stroke
0.76 0.76 0.74 0.76 0.75
0.78 0.80 0.77 0.79 0.78
Ulcerative colitis
0.75 0.75 0.68 0.70 0.76
0.78 0.80 0.48 0.68 0.55
Osteoarthritis
0.77 0.77 0.78 0.77 0.77
0.76 0.76 0.75 0.76 0.77
Ischemic stroke
0.77 0.77 0.76 0.72 0.76
0.79 0.79 0.75 0.72 0.77
Macular degeneration
0.77 0.77 0.75 0.68 0.78
0.82 0.77 0.66 0.60 0.81
Asthma
0.61 0.61 0.60 0.60 0.61
0.69 0.69 0.67 0.70 0.70
−0.4 −0.2 0.0 0.2
Change in AUROC
−0.4 −0.2 0.0 0.2
Change in AUROC
L1 LogReg
L2 LogReg
LGBM
MLP
Transformer
← Higher in UKB
Higher in AoU →
← Higher in UKB
Higher in FG →

**Figure S3. Prevalent classification model performance for all architectures**

We train classification (prevalent) models for all diseases in the UKB across 5 architectures: L1 penalized (LASSO) logistic regression, L2 penalized (ridge) logistic regression, light gradient boosting machines (LGBM), multilayer perceptron (MLP), and a fine-tuned transformer. AUROC for each architecture and 95% confidence interval for the change in AUROC between AoU (left) and FinnGen (right).

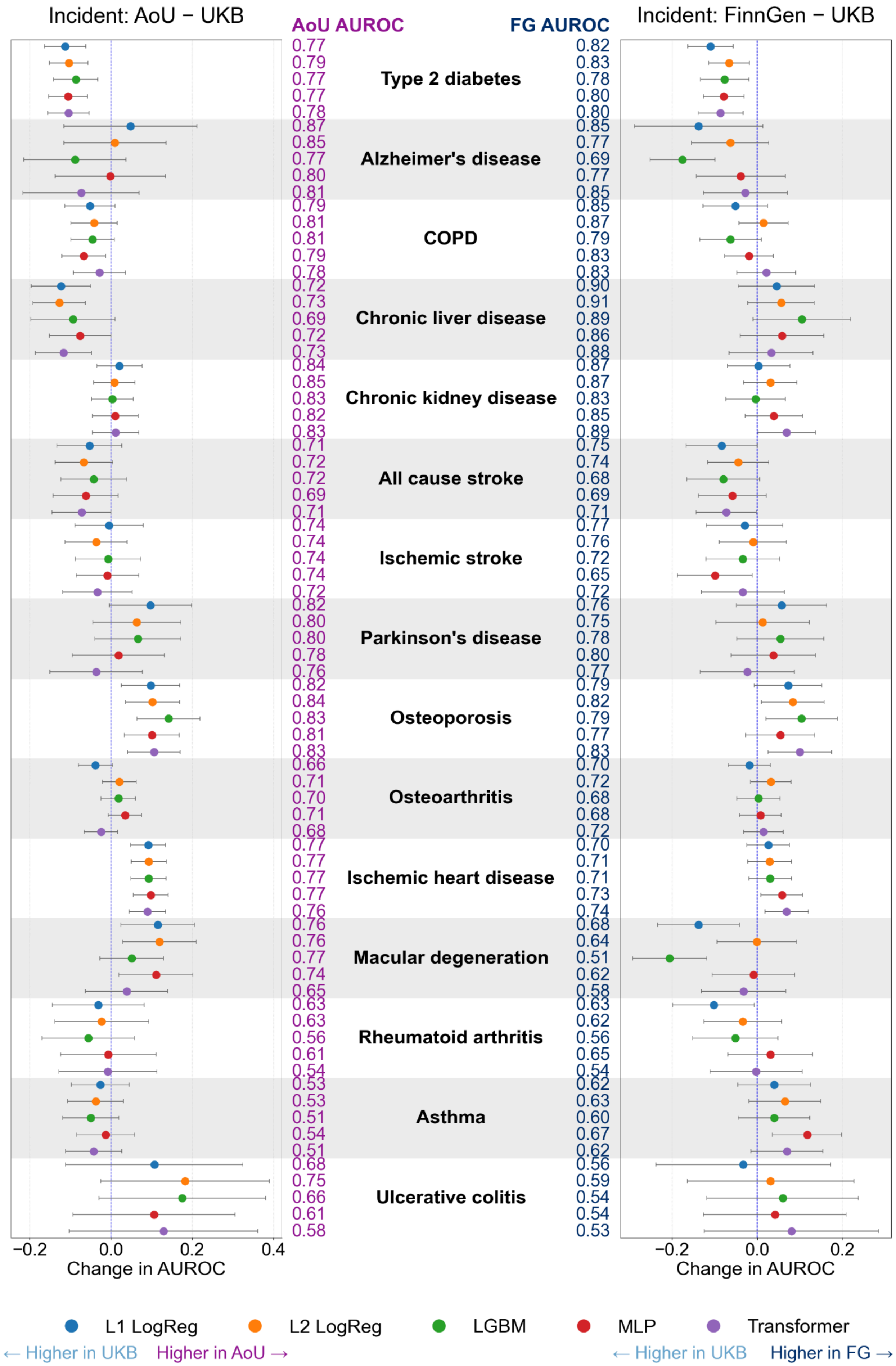

Incident: AoU − UKB
AoU AUROC
FG AUROC
Incident: FinnGen − UKB
Type 2 diabetes
0.77 0.79 0.77 0.77 0.78
0.82 0.83 0.78 0.80 0.80
Alzheimer's disease
0.87 0.85 0.77 0.80 0.81
0.85 0.77 0.69 0.77 0.85
COPD
0.79 0.81 0.81 0.79 0.78
0.85 0.87 0.79 0.83 0.83
Chronic liver disease
0.72 0.73 0.69 0.72 0.73
0.90 0.91 0.89 0.86 0.88
Chronic kidney disease
0.84 0.85 0.83 0.82 0.83
0.87 0.87 0.83 0.85 0.89
All cause stroke
0.71 0.72 0.72 0.69 0.71
0.75 0.74 0.68 0.69 0.71
Ischemic stroke
0.74 0.74 0.74 0.74 0.72
0.77 0.76 0.72 0.65 0.72
Parkinson's disease
0.82 0.80 0.80 0.78 0.76
0.76 0.75 0.78 0.80 0.77
Osteoporosis
0.82 0.84 0.83 0.81 0.83
0.79 0.82 0.79 0.77 0.83
Osteoarthritis
0.66 0.71 0.70 0.71 0.68
0.70 0.72 0.68 0.68 0.72
Ischemic heart disease
0.77 0.77 0.77 0.77 0.76
0.70 0.71 0.71 0.73 0.74
Macular degeneration
0.76 0.76 0.77 0.74 0.65
0.68 0.64 0.51 0.62 0.58
Rheumatoid arthritis
0.63 0.63 0.56 0.61 0.54
0.63 0.62 0.56 0.65 0.54
Asthma
0.53 0.53 0.51 0.54 0.51
0.62 0.63 0.60 0.67 0.62
Ulcerative colitis
0.68 0.75 0.66 0.61 0.58
0.56 0.59 0.54 0.54 0.53
−0.2 0.0 0.2 0.4
Change in AUROC
−0.2 0.0 0.2
Change in AUROC
L1 LogReg
L2 LogReg
LGBM
MLP
Transformer
← Higher in UKB
Higher in AoU →
← Higher in UKB
Higher in FG →

**Figure S4. Incident prediction model performance for all architectures**
We train prediction (incident) models for all diseases in the UKB across 5 architectures: L1 penalized (LASSO) logistic regression, L2 penalized (ridge) logistic regression, light gradient boosting machines (LGBM), multilayer perceptron (MLP), and a fine-tuned transformer. AUROC for each architecture and 95% confidence interval for the change in AUROC between AoU (left) and FinnGen (right).

The main metric that we consider for evaluating models across cohorts is the AUROC, but we also evaluate the partial AUROC up to a maximum false positive rate of 5%. While AUROC is a useful metric for determining model performance, it does not completely describe prediction specificity. We find no differences in average partial AUROC between UKB and FinnGen ($t = 0.81$, $p = 0.43$) for prevalent classification models but average partial AUROC in AoU is decreased ($t = 2.56$, $p = 0.024$). Several individual diseases show decreases in generalizability when considering partial AUROC (Fig S5). For prevalent type 2 diabetes, the partial AUROC does not generalize to both FinnGen and AoU, consistent with results for the overall AUROC. Prevalent Parkinson's, chronic liver disease, and asthma models also do not generalize to AoU when considering partial AUROC, again consistent with results for overall AUROC. However, COPD, rheumatoid arthritis, and osteoarthritis show new decreases in generalizability to AoU when considering partial AUROC. The osteoarthritis model also decreases in generalizability to FinnGen for partial AUROC. For incident disease prediction, we find no difference in average partial AUROC compared to UKB in AoU ($t = 0.95$, $p = 0.36$) and FinnGen ($t = 0.53$, $p = 0.60$). As above, the type 2 diabetes prediction models do not generalize for partial AUROC to both AoU and FinnGen, consistent with AUROC results. Furthermore, chronic liver disease prediction does not generalize to AoU, consistent with findings for AUROC. There is also a new decrease in generalizability for COPD in AoU for partial AUROC.

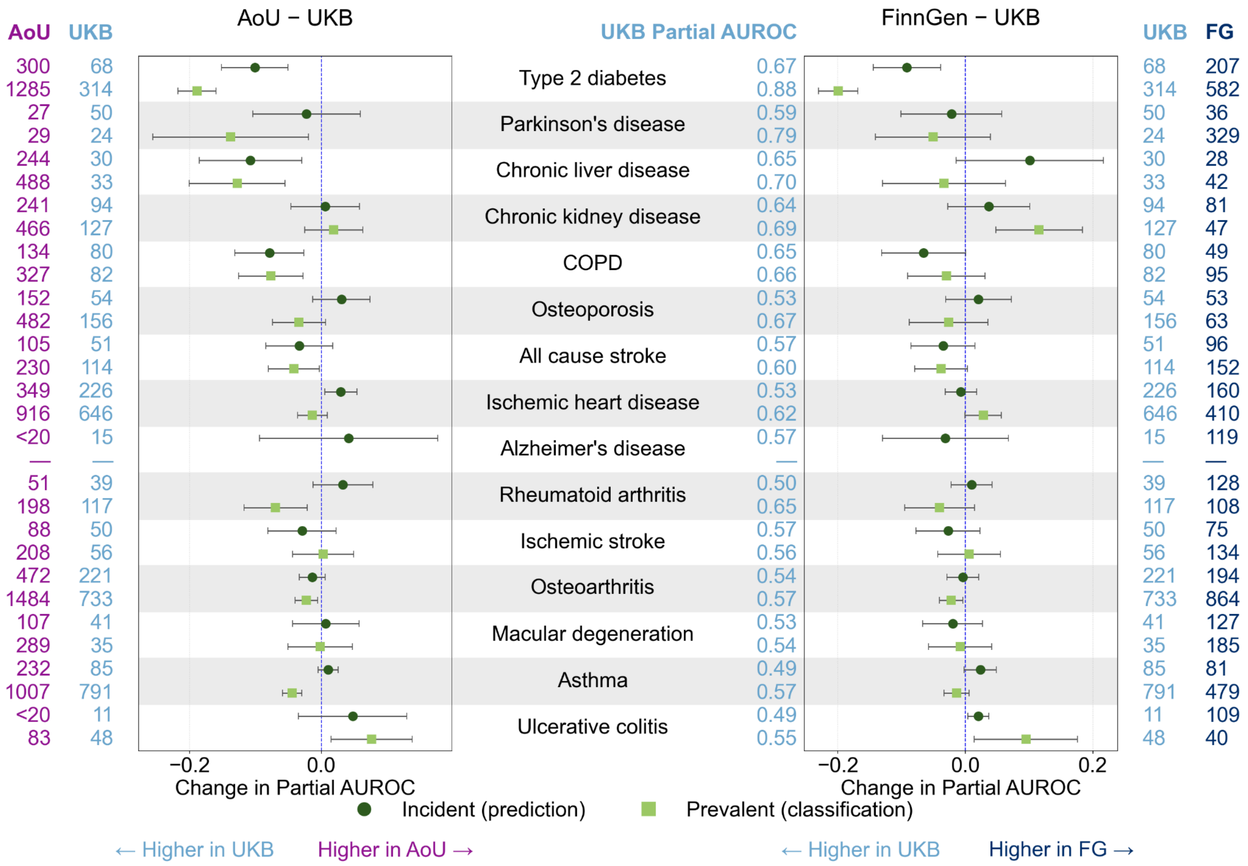


**Figure S5. Partial AUROC external cohorts**
External validation of UKB-trained incident (top row, green circle) and prevalent (bottom row, green square) proteomics disease classification models in FinnGen and AoU evaluated using partial AUROC with a maximum 5% false positive rate. Change in partial AUROC per disease between UKB and each external test set (AoU left, FinnGen right) for the ridge regression architecture with 95% confidence intervals, ordered by average UKB performance across prevalent and incident. Change < 0 indicates better performance in UKB while change > 0 indicates better performance in the external dataset. Case counts for each dataset are shown on the outer columns and UKB holdout partial AUROC values are in the center.

### Factors influencing generalizability

A variety of participant characteristics including age, genetic ancestry, BMI, and smoking status as well as phenotype quality may influence the generalizability of disease models across cohorts. In the main text we discuss that requiring two diagnosis codes in All of Us improves model performance for all diseases. In FinnGen, requiring two codes has much less of an effect on resultant case counts. For example, for prevalent type 2 diabetes, 8 cases are removed in FinnGen while 138 are removed in AoU; for incident type 2 diabetes, 4 cases are removed in FinnGen while 103 cases are removed in AoU. Furthermore, we perform a sensitivity analysis of type 2 diabetes model performance when UKB diabetes disease status is coded using only ICD10 and unambiguous ICD9 codes, compared to the comprehensive disease definition described thus far. We find similar UKB hold out performance (ΔAUROC prevalent = 0.0, ΔAUROC incident = 0.03). We also find similar performance when applying these new models to AoU (ΔAUROC = 0.0 for prevalent and incident) and FinnGen (ΔAUROC prevalent = 0.02, ΔAUROC incident = 0.0), comparing the AUROC for the model trained only on ICD codes vs the original coding design.

As the All of Us cohort selected for maximal ancestral diversity of participants, we use this cohort to further evaluate the impact that demographics and other variables such as BMI and genetic ancestry may have on model performance by stratifying the proteomics cohort. For age, we compare model performance in the subset of individuals below vs above the median cohort age, for genetic ancestry we require at least 1000 participants per group for prevalent disease and 500 for incident disease, for BMI we compare those below versus above median cohort BMI, and finally we compare smokers vs non smokers, where smokers are defined as individuals with at least 100 lifetime cigarettes. For all participant characteristics we additionally require at least 20 cases per group. We find no consistent differences in model performance stratified by participant characteristics across disease, for both classification (Fig. S6) and prediction (Fig. S7) models. We further evaluate performance across all ancestry groups regardless of group size for the type 2 diabetes models (Fig S8). We see that performance across non-EUR ancestry groups varies.

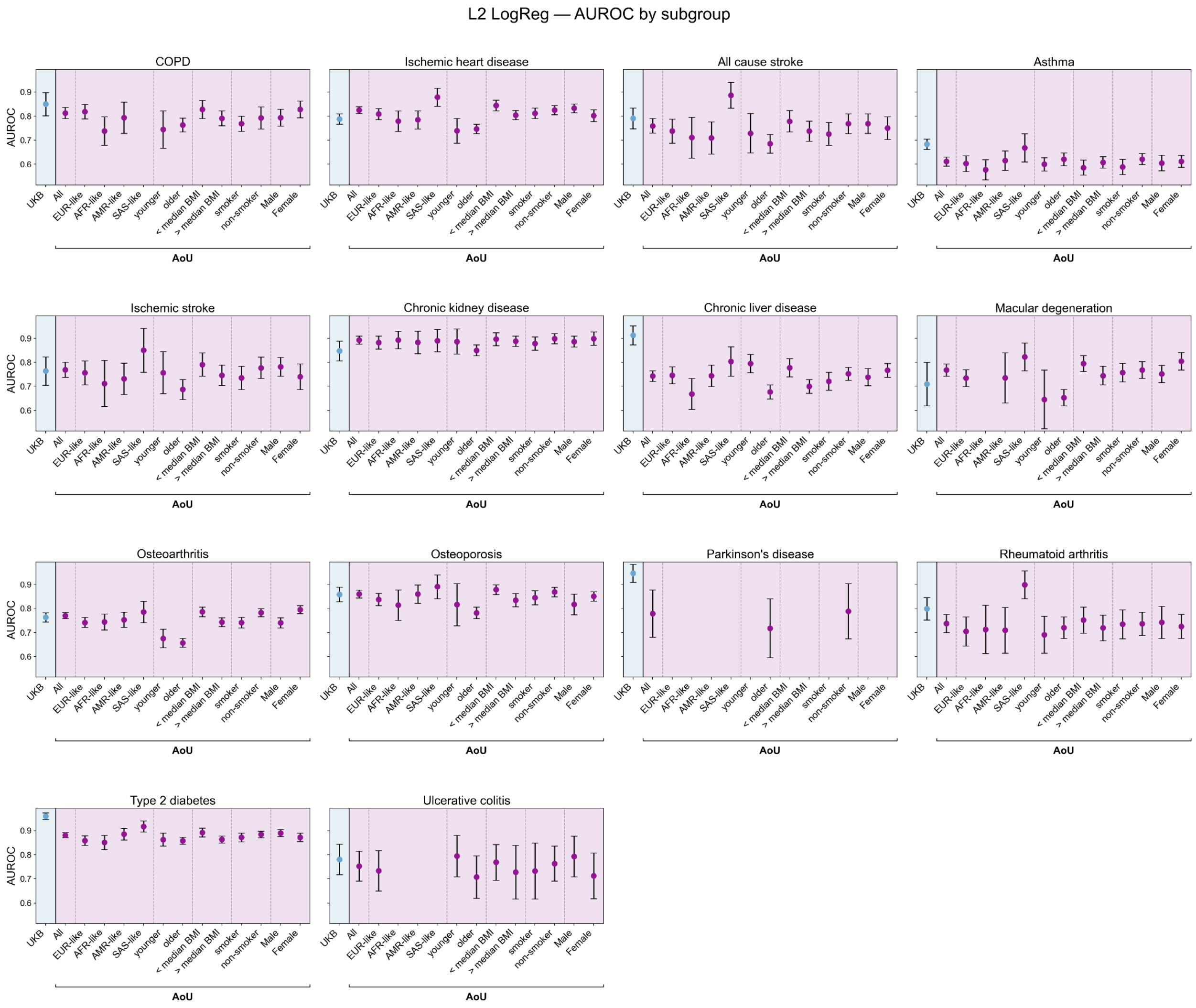


**Figure S6. AoU classification performance stratified by participant characteristics**

We stratify prevalent model performance in the AoU cohort by age (younger defined as age < median age and older defined as age >= median age), genetic ancestry (requiring at least 1,000 participants per group for prevalent disease and 500 for incident disease), BMI (above vs below median cohort BMI), smoker (at least 100 lifetime cigarettes) vs non smoker, and sex for all diseases. Groups with <20 cases are not shown.

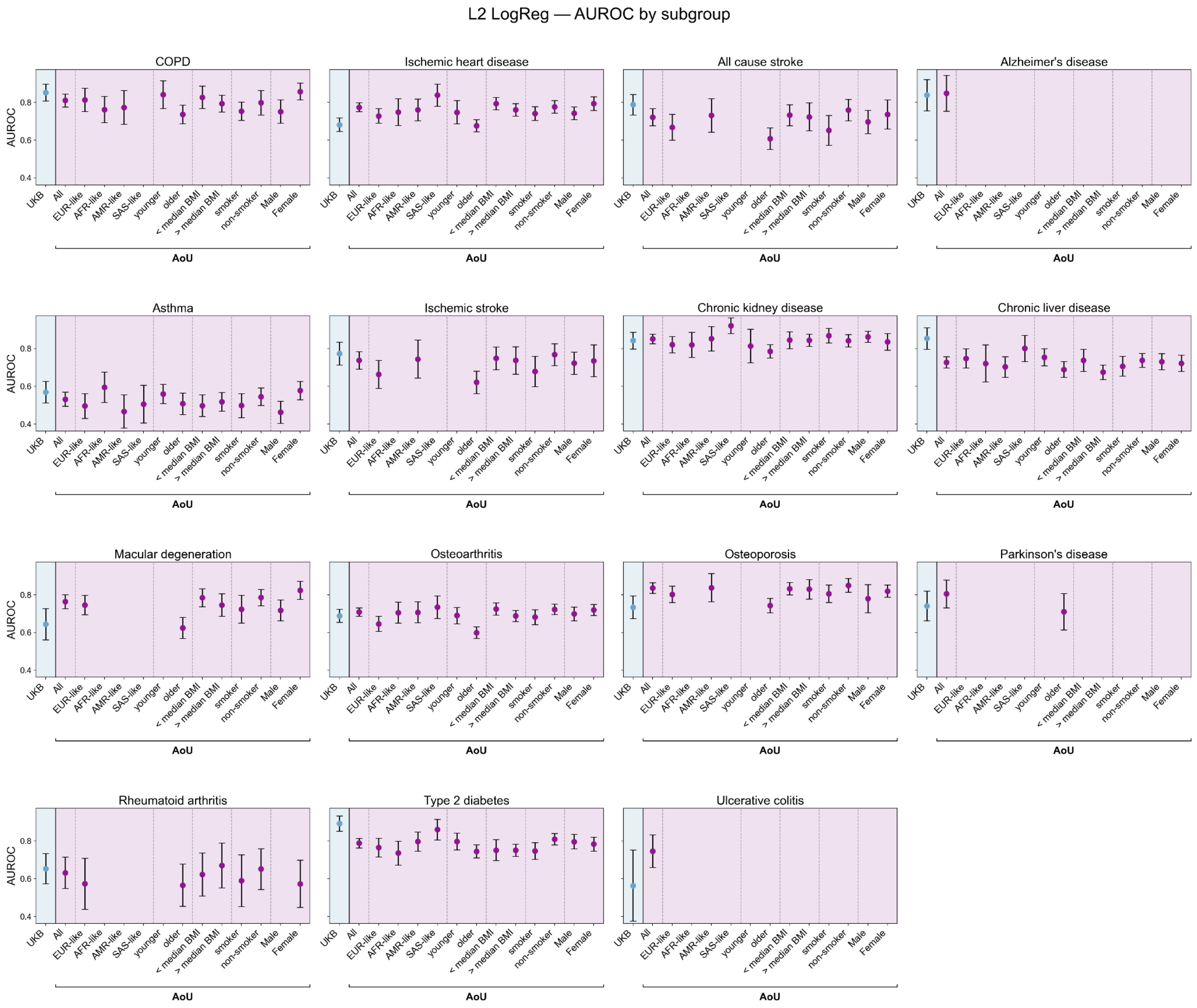


**Figure S7. AoU prediction performance stratified by participant characteristics**

We stratify incident model performance in the AoU cohort by age (younger defined as age < median age and older defined as age >= median age), genetic ancestry (requiring at least 1,000 participants per group for prevalent disease and 500 for incident disease), BMI (above vs below median cohort BMI), smoker (at least 100 lifetime cigarettes) vs non smoker, and sex for all diseases. Groups with <20 cases are not shown.

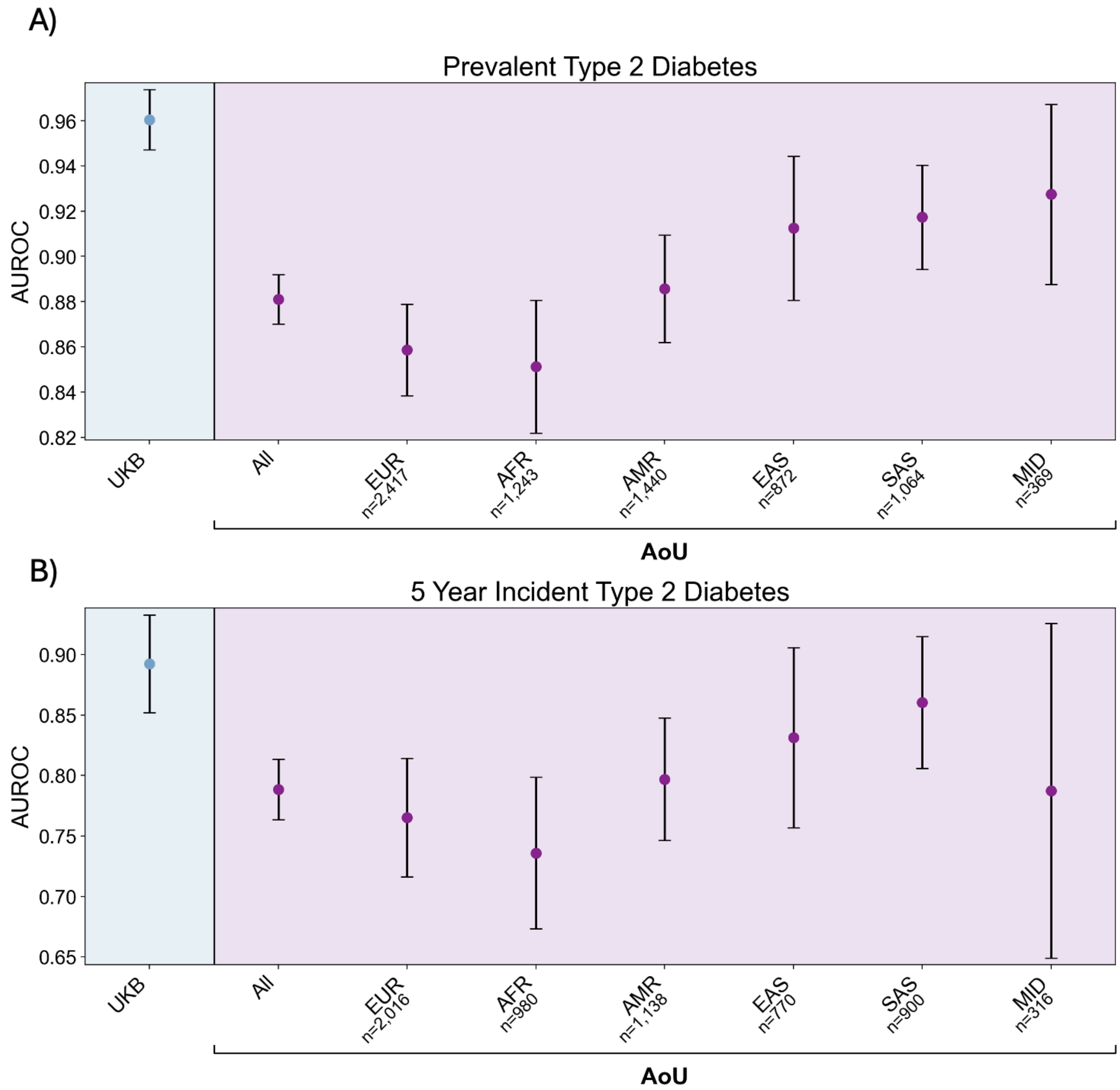


**Figure S8. AoU prediction performance stratified by genetic ancestry**
We stratify type 2 diabetes model performance in the AoU cohort by the full set of inferred genetic ancestry groups available in AoU for a) prevalent and b) incident disease.

To further understand type 2 diabetes model performance in AoU, we train prevalent and incident type 2 diabetes models on AoU proteomics data using a 90%:10% train:test split. We then evaluate both the AoU trained model and the UKB trained model on the same 10% hold out AoU set and calculate the AUROC (Fig4C-D) and AUPRC (Fig S9A-B). We also compare the predictive power of the Olink Explore HT panel, with 5,415 proteins, to the subset of proteins that overlaps with the Olink Explore 3072 panel, with only 2,920 proteins. We train models to classify and predict type 2 diabetes in AoU using each panel, with the same train/test split. We evaluate performance using AUROC and AUPRC. We find that the 5k panel does not outperform the 3k panel in any case (Fig. S9C-F).

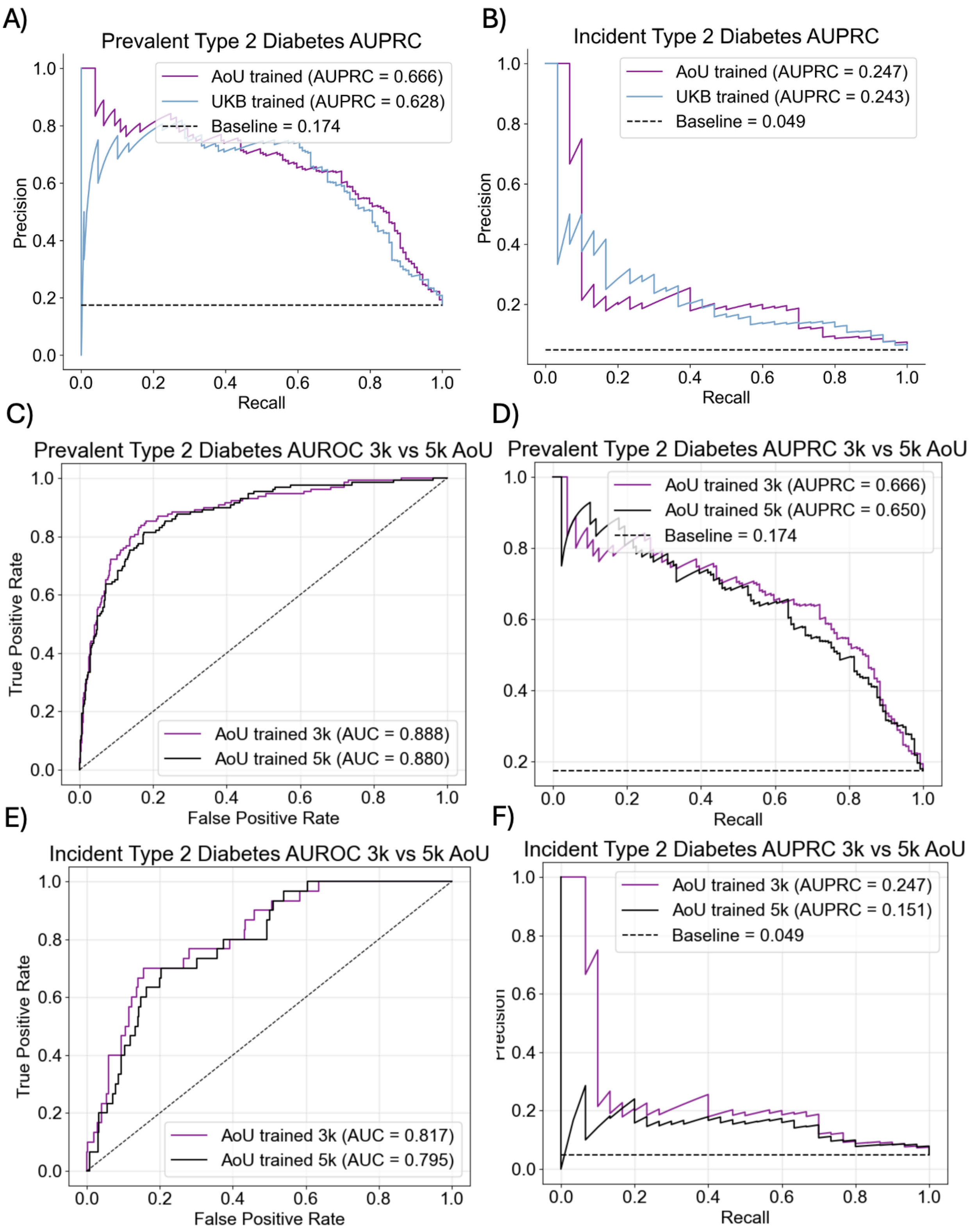


**Figure S9. AoU type 2 diabetes prediction**

We train a ridge regression to predict A) prevalent and B) incident type 2 diabetes using a 90%:10% training testing split in AoU and evaluate the resultant AUPRC. We then compare model performance when the dataset is just the subset of proteins included in UKB (3k panel) versus the full set of proteins included in AoU (5k panel) for prevalent disease C) AUROC and D) AUPRC and incident disease E) AUROC, and F) AUPRC.

Technical factors such as the Olink proteomic platform may also affect generalizability. To assess this, we apply our model to each of two sub-cohorts in FinnGen, evaluating Cohort A which uses the same Olink 3072 platform as UKB and Cohort B which uses the Olink Explore HT panel. We find that most diseases generalize across subcohorts, with the exception of prevalent and incident type 2 diabetes in cohort B, and incident all cause stroke cohort A (Fig S10). While the diabetes models do generalize in cohort A and not cohort B, we do not believe this reflects a difference due to the proteomic platform. First, if the proteomic platform was significantly affecting model performance we would expect systematic shifts in generalizability between the two FinnGen cohorts. Instead, we see that overall most models generalize in both cases. Comparing the T2D ridge regression coefficients of proteins missing from each external panel relative to UKB, proteins absent from external cohort panels account for only a small percent of the model's total absolute coefficient mass (0.6% FinnGen Cohort A prevalent, 0.85% FinnGen Cohort A incident, 2.3% FinnGen Cohort B prevalent, 2.7% FinnGen Cohort B incident, 2.4% AoU prevalent, 2.8 % AoU incident). This is a small differential relative to the observed AUROC gap (0.058 FinnGen Cohort B prevalent, 0.064 FinnGen Cohort B incident, 0.079 AoU prevalent, 0.10 AoU incident). Additionally, none of the top 20 highest-weighted proteins in the prevalent T2D model are missing from any external cohort. In addition, ADIPOQ is the top-ranked protein in the incident T2D ridge regression model and is absent from all three external cohorts including Cohort A, indicating the model is robust to the loss of even its most heavily-weighted feature and that panel content differences alone cannot explain the Cohort B generalization drop. Overall, this suggests that the Olink platform does not have a strong effect on model generalizability.

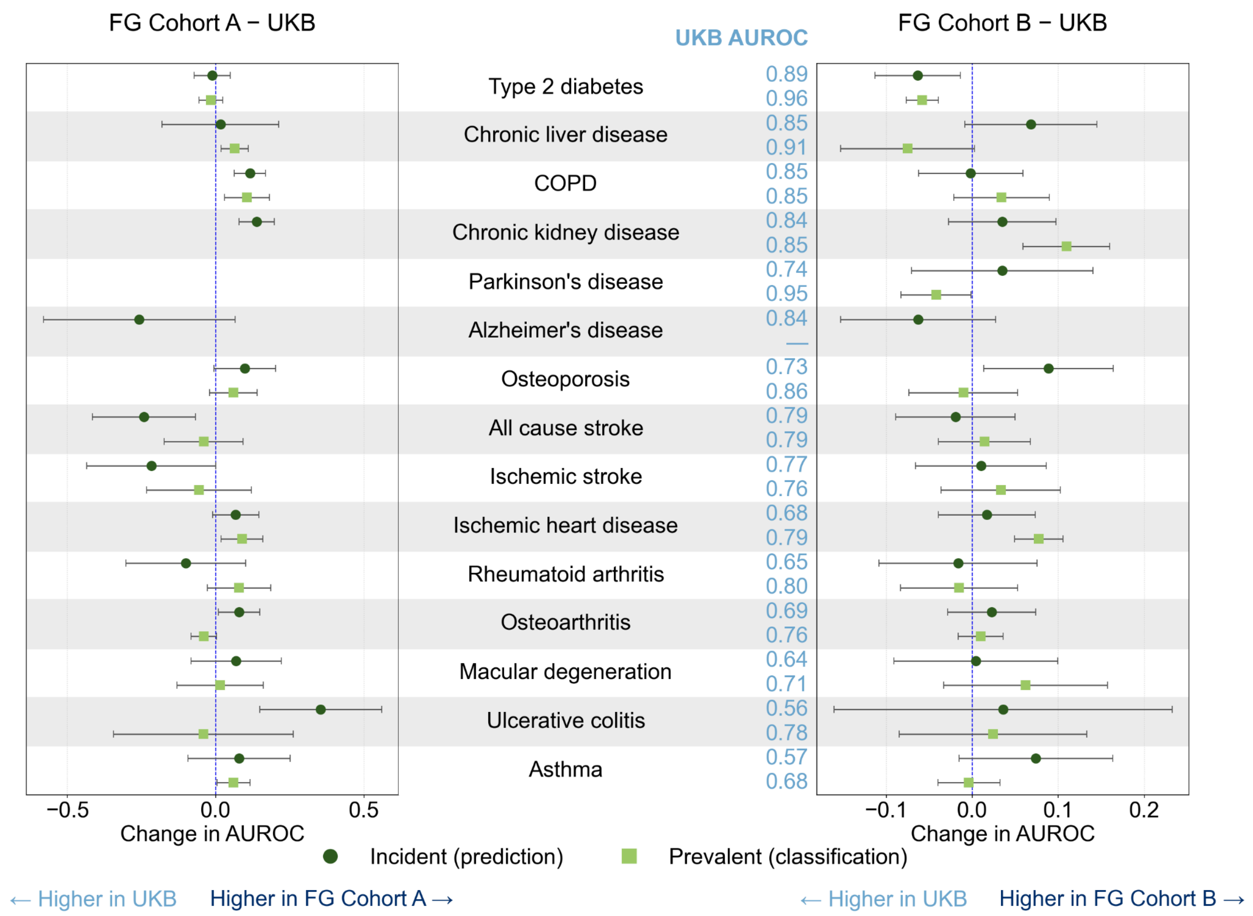


**Figure S10. FinnGen subcohort performance**
External validation of UKB-trained incident (top row, green circle) and prevalent (bottom row, green square) ridge regression proteomics disease classification models in FinnGen Cohort A (n=1925, Olink Explore 3072) and Cohort B (n=3940, Olink Explore HT) compared to hold out data in UKB (n=5303, Olink Explore 3072). Change in AUROC per disease between UKB and each external test set (Cohort A left, Cohort B right) for the ridge regression architecture with 95% confidence intervals, ordered by UKB performance. Change < 0 indicates better performance in UKB while change > 0 indicates better performance in the external dataset. Case counts for each dataset are shown on the outer columns and UKB holdout AUROC values are in the center.

### UKB cohort structure

To further understand the proteomics data structure in the UKB cohort we run principal component analysis (PCA) on the full cohort. We use the raw data and normalize data using a standard scalar (mean 0, standard deviation 1). We then plot principal component 1 (PC1) vs PC2 and evaluate how well these two components stratify the data in terms of age, sex, BMI, and smoking status (Figure S11). Individuals missing a given participant characteristic are excluded from the relevant plot. We find that the first two PCs do not provide clear clustering of any of these characteristics. Next, we fit a PCA on only the training set described earlier for proteomic modeling. We then train a random forest predictor using 500 estimators from the top 20 PCs for age, sex, BMI, and smoking status, along with prevalent and incident type 2 diabetes on this same training set. Finally, we apply both the PCA and the random forest to the hold out test set. We plot the two PCs that contribute the most to each random forest, colored by outcome variable for the hold out test set (Figure S12). The random forest achieves AUROC 0.997 for sex, 0.658 for smoking status, 0.843 for prevalent diabetes, and 0.745 for incident diabetes. Performance for all diseases can be found in Table S8. The mean absolute error is 4.2 for age and 2.5 for BMI.

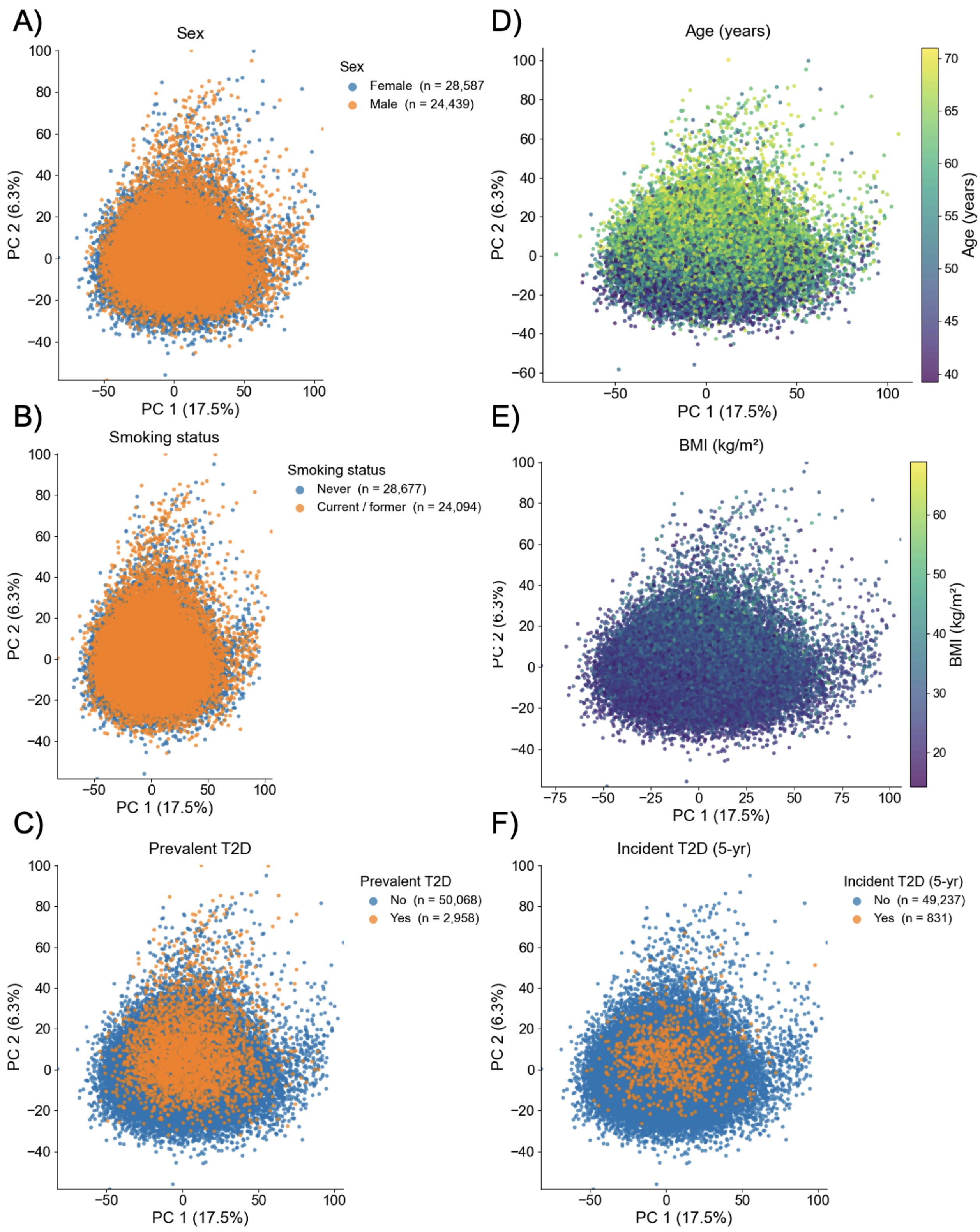


**Figure S11. PCA of UKB proteomics**

We run principal component analysis (PCA) on the full cohort of UKB proteomics data and plot the first two principal components, PC1 and PC2, colored by a) sex b) smoking status c) prevalent type 2 diabetes case status d) age e) BMI and f) incident type 2 diabetes case status. Number of individuals in each group is shown in the legend.

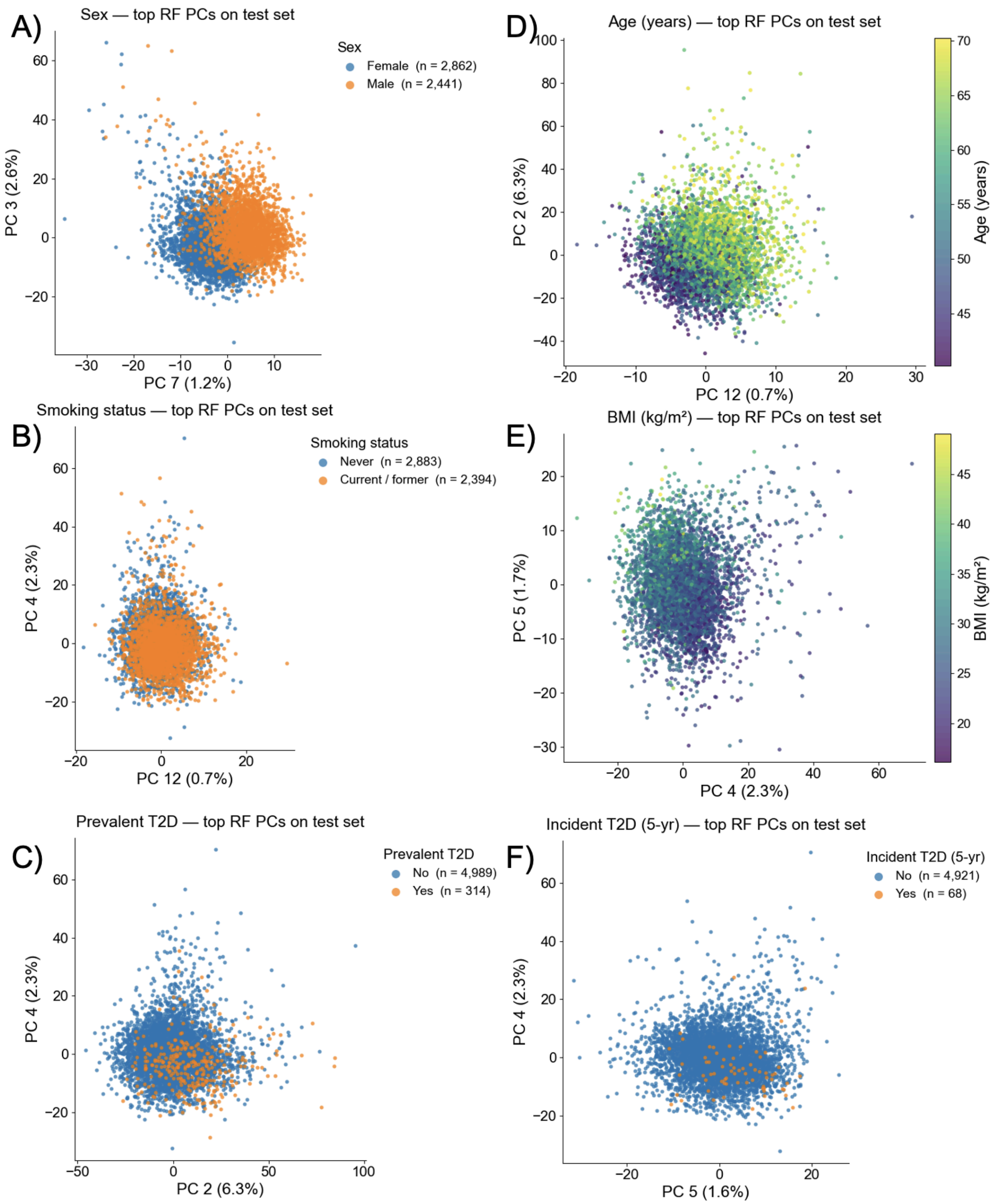

**Figure S12. Random forest from UKB principal components**
Using our proteomics training set we fit a new PCA using only the training set. We then train a random forest from the first 20 PCs to predict a) sex b) smoking status c) prevalent type 2 diabetes case status d) age e) BMI and f) incident type 2 diabetes case status. Using the random forest feature importance, we calculate the top two PCs for each model and plot these two PCs in the test set, colored by target variable.

### Proteomic univariate analysis

We conduct univariate association analysis for each protein (n=2,920) for each of the prevalent and incident diseases in the UKB. Each protein was standardized to zero mean and unit variance (z-score) across participants. All models were adjusted for age at baseline and self-reported sex. Age was z-scored across the cohort; sex was modeled as a categorical variable. For prevalent disease, we estimate association between proteins and disease using a logistic regression model. For 5 year incident disease, we fit a Cox proportional hazards model using lifelines (CoxPHFitter) on each protein–disease pair, regressing time to disease event. For all associations, p values are adjusted within each disease using Benjamini–Hochberg false discovery rate with alpha 0.05. We then calculate a vector of the regression coefficients across the proteome for each disease (β vector). Coefficients with an adjusted p value < 0.01 are set to 0. To evaluate similarity in the proteome across diseases we compute the correlation of this β vector across diseases, compared to the correlation of phenotypes directly (Fig. S13A).

### Model interpretation

We apply disease models to predict all other diseases to evaluate model specificity. For incident model results see main text (Fig 5). For prevalent classification, the models are largely most predictive of the disease they are trained on, with some exceptions (e.g. ischemic stroke -> Parkinson's); however, each disease is best predicted by the model for which it is trained (Fig S13B).

A)

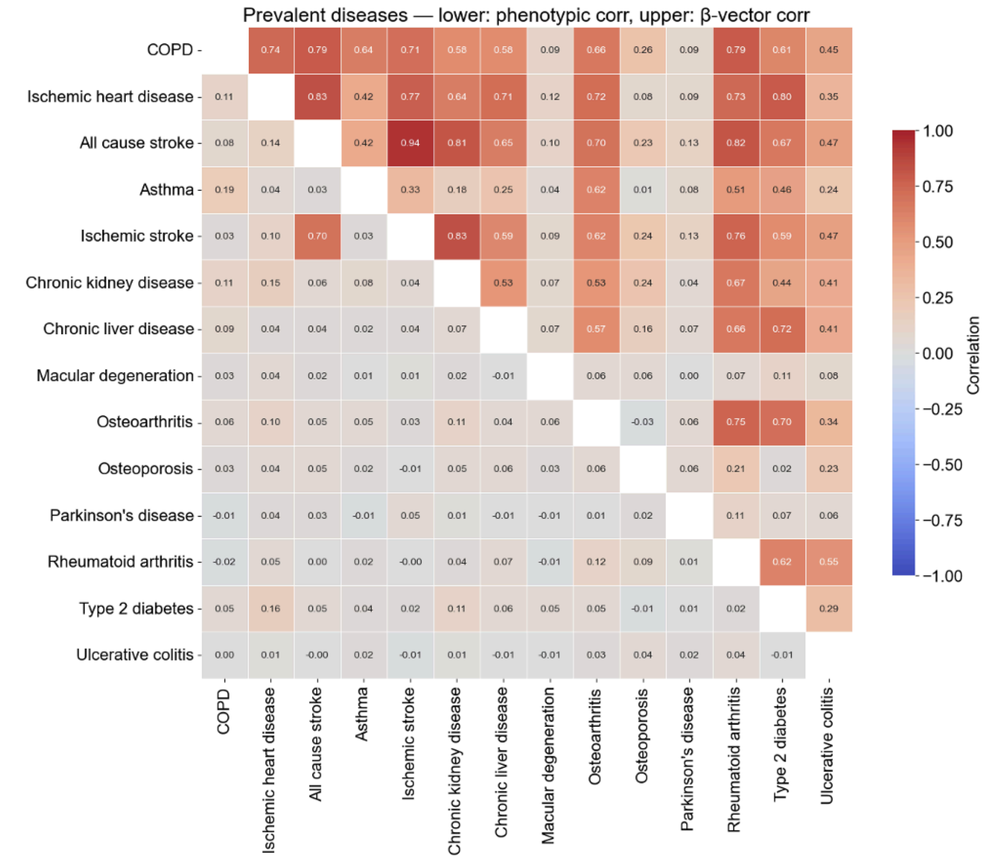

Prevalent diseases — lower: phenotypic corr, upper: β-vector corr
COPD
Ischemic heart disease
All cause stroke
Asthma
Ischemic stroke
Chronic kidney disease
Chronic liver disease
Macular degeneration
Osteoarthritis
Osteoporosis
Parkinson's disease
Rheumatoid arthritis
Type 2 diabetes
Ulcerative colitis
Correlation


B)

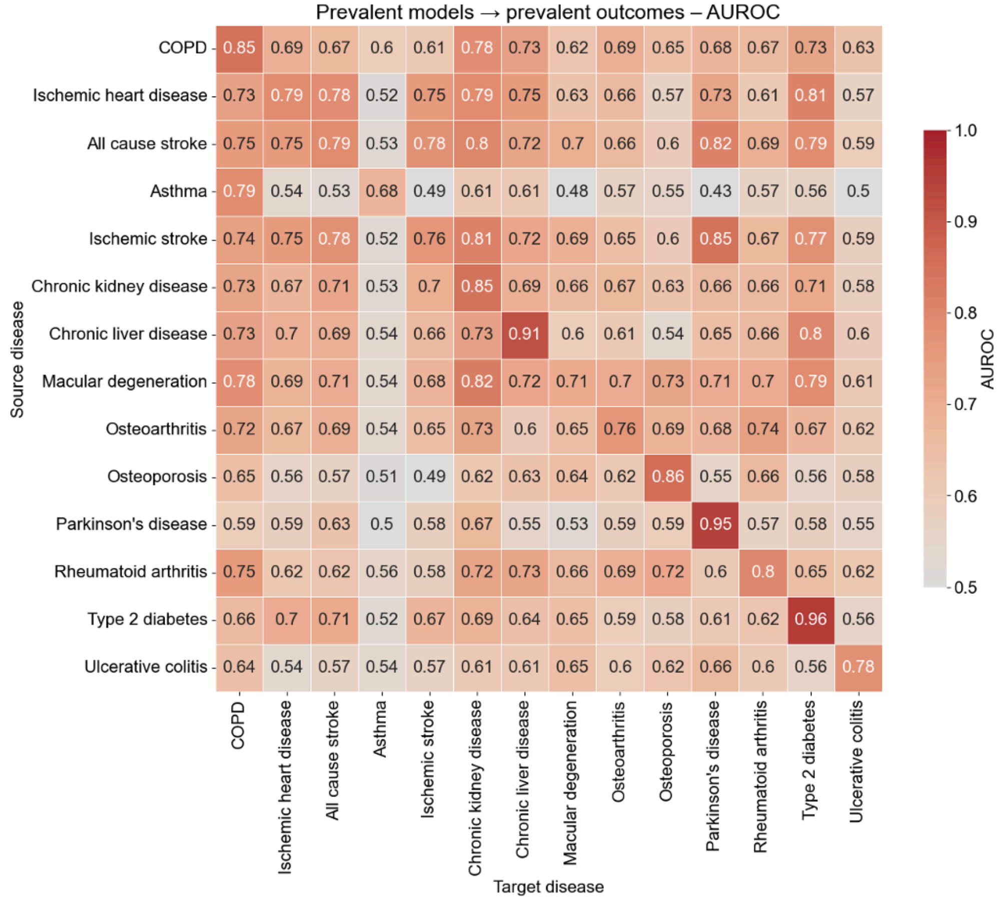

Prevalent models → prevalent outcomes – AUROC
Source disease
Target disease
COPD
Ischemic heart disease
All cause stroke
Asthma
Ischemic stroke
Chronic kidney disease
Chronic liver disease
Macular degeneration
Osteoarthritis
Osteoporosis
Parkinson's disease
Rheumatoid arthritis
Type 2 diabetes
Ulcerative colitis
AUROC

**Figure S13. Prevalent model specificity**

A) Combined correlation heatmaps for prevalent diseases. Pearson correlations between binary disease indicators (phenotypic correlation, lower triangle) and disease-specific coefficient vectors derived from univariate proteomic association analyses (β-vector correlation, upper triangle). For each disease, coefficient vectors span 2,920 plasma proteins from the UK Biobank Olink panel; coefficients with an adjusted p-value ≥ 0.01 were set to zero prior to correlation.

B) Performance of L2 ridge regression models applied to all other diseases and evaluated using AUROC for prevalent diseases. Source disease is the disease used to evaluate and target disease is the disease being predicted by the source model.

In addition, we perform a PheWAS for each proteomic disease score. We calculate the Pearson correlation and Bonferroni adjusted p value between the model score and 11,449 phenotypes in UKB. We find that the top associations often show domain specificity. For example, HbA1c and BMI are among the top 10 associations in the incident type 2 diabetes PheWAS (Fig S14).

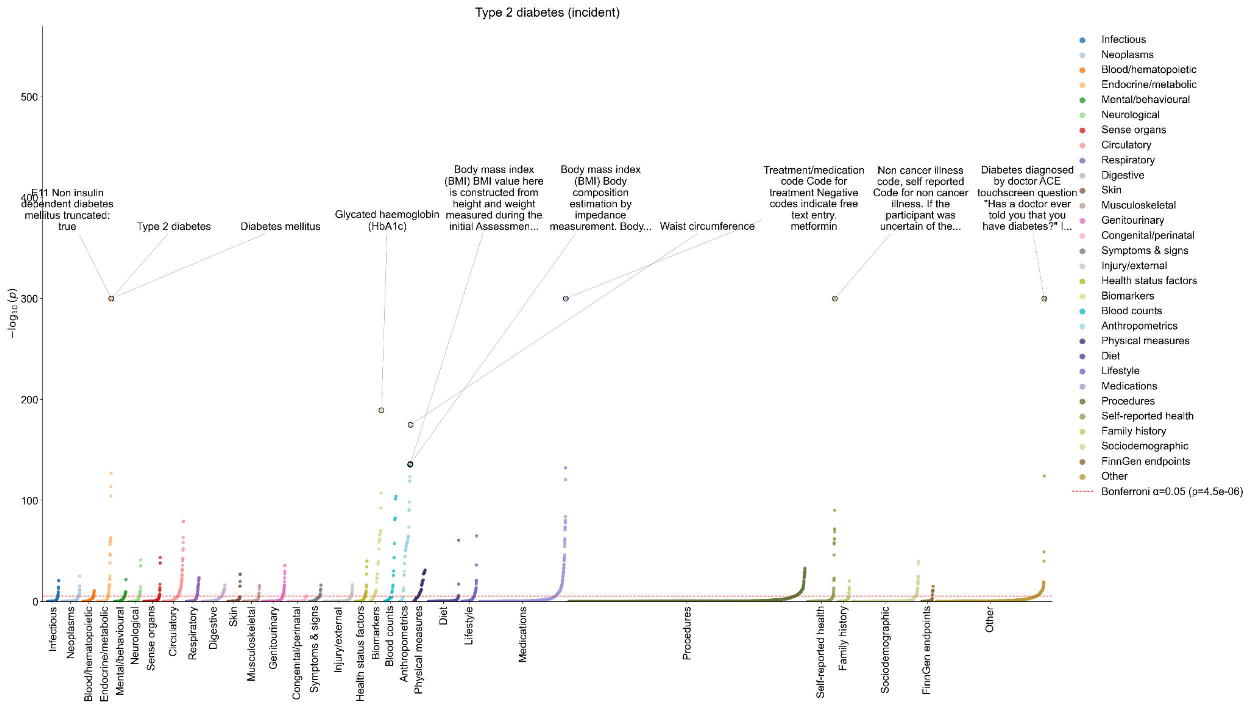


**Figure S14. PheWAS of incident type 2 diabetes**

Each point represents the association between the incident type 2 diabetes proteomic disease score and one UKB phenotype, plotted as $-\log_{10}$(nominal P) on the y-axis. Associations are calculated using pearson correlation. Phenotypes are grouped along the x-axis into 30 trait categories. Within each category, phenotypes are ordered by increasing significance. The red dashed line marks the Bonferroni-corrected threshold at $\alpha = 0.05$ ($P = 4.5 \times 10^{-6}$); points above it are phenome-wide-corrected significant. The associations with the ten lowest p values are labeled. Six phenotypes have p-values reported as zero (numerical underflow) and are plotted at $-\log_{10}(P) = 300$.

Genome-wide association analyses of protein scores

Genome-wide association analyses were performed in unrelated UK Biobank participants of European genetic ancestry (*4*) using SAIGE, a generalized linear mixed-model framework that accounts for cryptic relatedness and population structure via a sparse genetic relationship matrix (*5*). Null models included age, sex, genotyping array, and genetic principal components as covariates. To maximize statistical power for downstream genetic analyses, protein scores were generated for all UK Biobank participants using a 10-fold cross-validation framework, in which L2-regularized logistic regression models predicting disease-specific plasma protein signatures from Olink proteomic measurements were fit iteratively across 10 folds, yielding an out-of-fold predicted protein score for every participant. Separate models were constructed for incident and prevalent disease endpoints. Prediction models were developed for 29 disease traits, and the resulting protein scores were used as quantitative phenotypes in genome-wide association analyses.

We performed heritability analysis using linkage disequilibrium score regression (LDSC) (*6*, *7*). Observed-scale SNP heritability ranged from $h^2$ = 0.047 (SE = 0.016) for the incident Parkinson disease score to $h^2$ = 0.139 (SE = 0.017) for the prevalent asthma score. LDSC intercepts were close to 1 across scores (range, 0.948-0.997), indicating little evidence of residual inflation from population stratification or cryptic relatedness.

Genetic correlation analysis

Publicly available genome-wide association summary statistics were obtained for 30 UK Biobank traits spanning adiposity, cardiovascular, metabolic, lipid, musculoskeletal, respiratory, renal, psychological, sociodemographic, and substance-use domains, generated by genome-wide association analyses of predominantly EUR-ancestry UK Biobank participants (*4*).

Genome-wide genetic correlations between genetically predicted protein scores and UK Biobank trait GWAS were estimated using LDSC with the UK Biobank European LD reference panel (*6*, *7*). Pairwise analyses were performed between each of the 29 protein score GWAS and each of the 30 UKB trait GWAS (870 pairwise tests). Statistical significance was assessed using Benjamini–Hochberg false discovery rate (FDR) correction across all pairwise comparisons.

Protein scores for cardiometabolic and respiratory diseases were broadly correlated with BMI, including incident type 2 diabetes ($r_g$ = 0.68, FDR = $5.6\times10^{-63}$), incident liver disease ($r_g$ = 0.64, FDR = $1.7\times10^{-58}$), prevalent COPD ($r_g$ = 0.62, FDR = $6.0\times10^{-53}$), incident COPD ($r_g$ = 0.54, FDR = $4.7\times10^{-50}$), incident osteoarthritis ($r_g$ = 0.71, FDR = $8.3\times10^{-39}$), and incident rheumatoid arthritis ($r_g$ = 0.60, FDR = $7.5\times10^{-35}$; prevalent $r_g$ = 0.60, FDR = $1.9\times10^{-28}$). The incident type 2 diabetes score was also positively correlated with type 2 diabetes itself ($r_g$ = 0.80, FDR = $2.4\times10^{-51}$), consistent with its intended disease specificity. Respiratory scores showed similar evidence of biological coherence, showing their strongest and most specific overlap with respiratory traits (mean $r_g$ = 0.35; 67% Bonferroni-significant): incident and prevalent COPD scores were genetically correlated with smoking initiation ($r_g$ = 0.46, FDR = $2.2\times10^{-27}$ and $r_g$ = 0.51, FDR = $5.9\times10^{-36}$, respectively) and with COPD itself ($r_g$ = 0.69, FDR = $3.9\times10^{-32}$ and $r_g$ = 0.75, FDR = $4.9\times10^{-31}$, respectively). Asthma scores showed a similar pattern, with the prevalent asthma score strongly correlated with asthma itself ($r_g$ = 0.60, FDR = $1.0\times10^{-31}$) and the incident asthma score correlated with smoking initiation ($r_g$ = 0.35, FDR = $3.0\times10^{-14}$). These results suggest that the protein scores recapitulate genetic liability shared across adiposity, smoking behaviour, cardiometabolic disease, and respiratory disease. We compare this GWAS signal with the signal from the PheWAS described earlier and find that they are largely consistent (Fig S15).

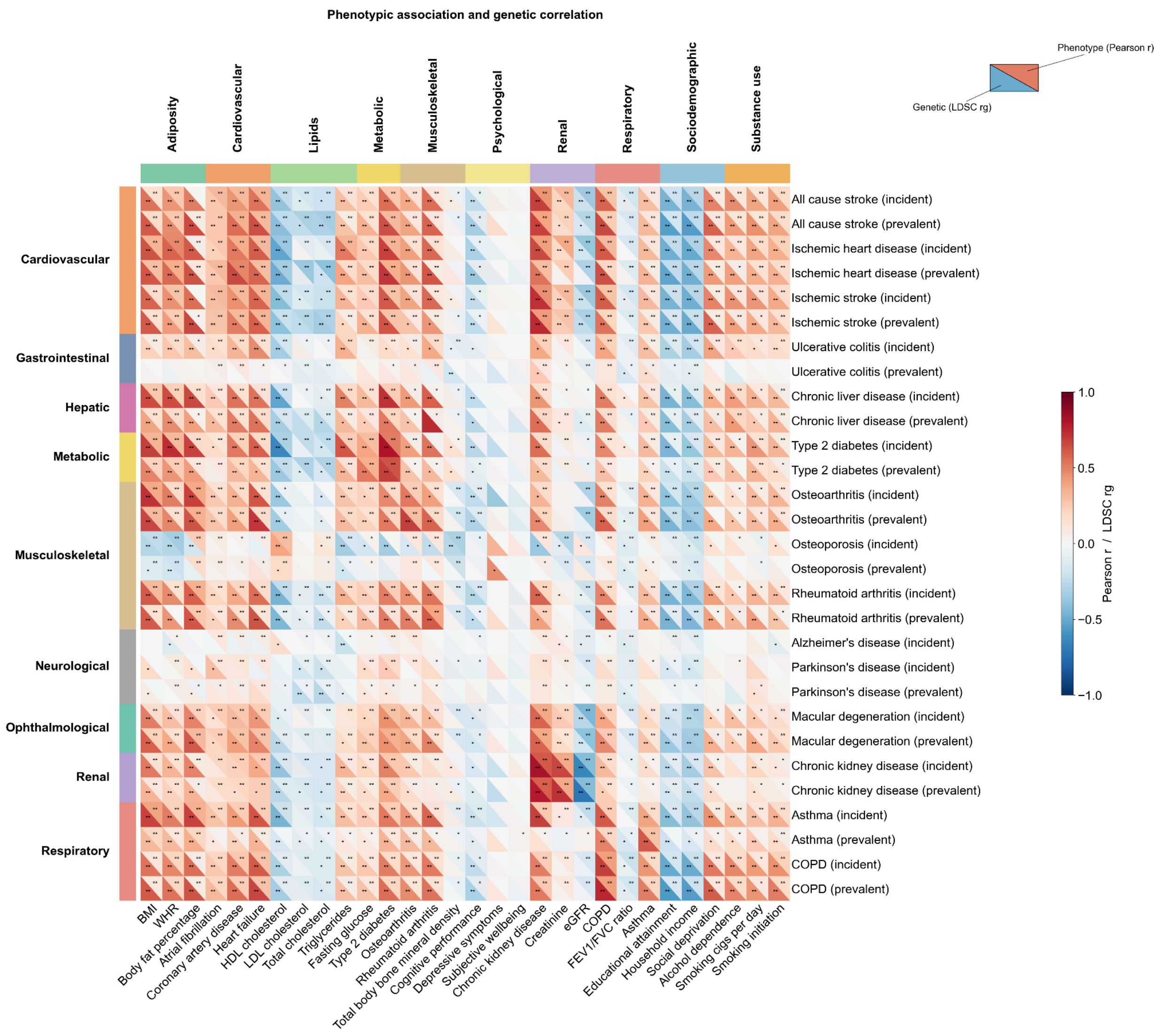


**Figure S15. Phenotype associations vs GWAS signal**
For 29 UK Biobank protein disease scores (rows) and 30 UK Biobank traits (columns), we computed both phenotypic Pearson correlations (top triangle) and genome-wide LDSC genetic correlations (rg, bottom triangle). Rows are grouped into 9 broad disease categories and columns into 10 UKB trait categories (colored side bars). For phenotypic association (top), we calculate the Pearson correlation between the proteomic disease model score and the UKB phenotype value. For genetic correlation (bottom), we computed genome-wide LDSC genetic correlations between the proteomic disease model score and the UKB phenotype value. For each disease category × trait category block, we estimated the mean rg and the percentage of Bonferroni-significant pairs to assess whether each score's genetic correlation profile is concentrated within its biologically corresponding domain or shared broadly across domains (full estimates for all blocks are provided in Supplementary Dataset 1). Cell color reflects the sign and magnitude of pearson r (top) or $r_g$ (bottom); asterisks denote significance (*nominal P < 0.05, **Bonferroni-corrected P < 0.05).

### Effect of training case count on model performance

The number of training cases varies in UKB across diseases, and case count also varies across cohorts. To evaluate the effect of training case count on model performance we downsample type 2 diabetes cases for prevalent and incident disease. We try two methods of downsampling: 1) downsampling cases only 2) downsampling cases and controls proportionally. We consider case counts of 50, 100, 250, 500, 1000, and 2500. We find that model performance on hold out test data consistently improves as more training cases are added for both prevalent and incident models (Fig S16). However, the prevalent models, which achieve higher performance, have diminishing increases in AUROC above 500 cases. There is a sharp increase in performance in both cases from 50 training cases to 100 training cases.

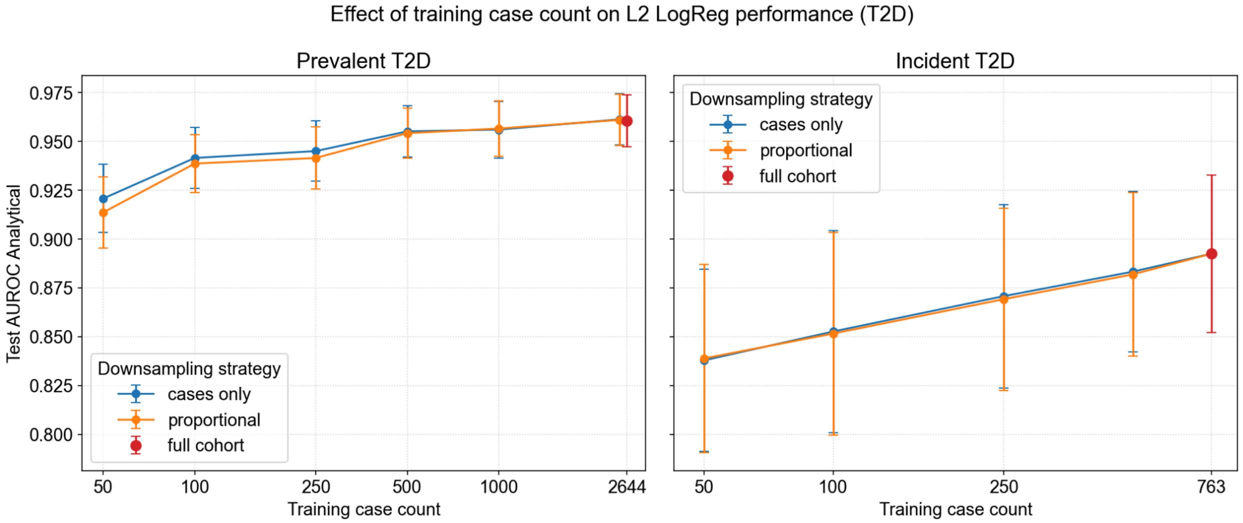


**Figure S16. Effect of training case count on type 2 diabetes model performance**
To evaluate the effect of training case count on model performance we downsample case counts by two methods 1) cases are downsampled to a fixed number of cases (cases only, blue) and 2) cases are downsampled to a fixed number and controls are also downsampled to maintain the same case/control ratio (proportional, orange). We train a new l2 logistic regression model for prevalent type 2 diabetes and incident type 2 diabetes using each downsampled training set and evaluate on the hold out test set. The right most point in each plot shows the test set performance using the full training cohort.

### Supplementary Tables

**Table S1. Curated disease phenotypes in UKB and AoU**

ICD10 and 9 codes were used for UK Biobank and All of Us, while UK Biobank also included baseline verbal interviews and measures. Asterisk indicates those phenotypes previously described (*2*).

| **Disease** | **ICD-10 codes (UKB + AoU)** | **ICD-9 codes (UKB+AoU)** | **Baseline Verbal Interview Diagnosis Codes (UKB only)** | **Baseline Measures (UKB only)** |
|---|---|---|---|---|
| Asthma | J45-J46 | 493 | 1111 | |
| Ulcerative Colitis | K51 | 556 | 1463 | |
| Type 2 Diabetes* | E11 | 250 | 1223 | Taking insulin medication (6153, 6177)<br>Non-fasting blood hbA1c $^{3}$ 48 mmol/mol (30750)<br>Non-fasting blood glucose $^{3}$ 11.1 mmol/L (30740) |
| Ischemic Heart Disease (IHD)* | I20-I25 | 410-414 | 1074, 1075 | |
| Ischemic Stroke | I63-I64 | 434,436 | 1583 | |
| All Stroke | I60-I64 | 430,431,434,436 | 1081, 1086, 1491, 1583 | |
| COPD* | J44 | 496 | 1112 | |
| Chronic Liver Disease* | K70, K73-K74, K75.8, K76.0 | 571 | 1157, 1158, 1604 | |
| Chronic Kidney Disease* | N18 | 585 | 1192, 1193, 1194 | |
| Alzheimer's* | F00, G30 | 331.0 | | |
| Parkinson's and parkinsonism* | G20-G22 | 332 | 1262 | |
| Rheumatoid Arthritis* | M05-M06 | 714 | 1464 | |
| Macular Degeneration* | H35.3 | 362.5 | 1528 | |
| Osteoporosis* | M80-M81 | 733.0 | 1309 | |
| Osteoarthritis* | M15-M19 | 715 | 1465 | |

#### Table S2. FinnGen phenotype mapping

| Disease | FinnGen Endpoints | Covered ICD codes |
|---|---|---|
| Asthma | J10_ASTHMA_EXMORE | Hospital discharge: ICD-10 — J45, J46<br>Hospital discharge: ICD-9 — 493<br>Hospital discharge: ICD-8 — 493<br>Cause of death: ICD-10 — J45, J46<br>Cause of death: ICD-9 — 493<br>Cause of death: ICD-8 — 493 |
| Ulcerative Colitis | K11_UC_STRICT2 | Hospital discharge: ICD-10 — K51<br>Hospital discharge: ICD-9 — 556<br>Hospital discharge: ICD-8 — 5631\|569<br>Hospital discharge: excluded ICD-9 — 5564A<br>Cause of death: ICD-10 — K51<br>Cause of death: ICD-9 — 556<br>Cause of death: ICD-8 — 5631\|569<br>Cause of death: excluded ICD-9 — 5564A |
| Type 2 Diabetes | T2D | Hospital discharge: ICD-10 — E11<br>Hospital discharge: ICD-9 — 250.A<br>Cause of death: ICD-10 — E11<br>Cause of death: ICD-9 — 250.A<br>Medicine purchases: ATC — A10B |
| Ischemic Heart Disease (IHD) | I9_IHD,I9_ANGINA,<br>I9_UAP,<br>I9_MI_STRICT,<br>I9_CORATHER | Hospital discharge: ICD-10 — I20-I25,T82.2, Z95.1<br>Hospital discharge: ICD-9 — 41[0-4]\|9960A<br>Hospital discharge: ICD-8 — 41[0-4]<br>Cause of death: ICD-10 — I20, I21, I22, I23, I24, I25, I46, R96, R98, T82.2, Z95.1<br>Cause of death: ICD-9 — 41[0-4]\|798\|9960A<br>Cause of death: ICD-8 — 41[0-4]\|798<br>Cause of death: excluded ICD-9 — 7980A<br>KELA reimbursements: KELA codes — 206 |
| Ischemic Stroke | I9_STR_EXH | Hospital discharge: ICD-10 — I63, I64<br>Hospital discharge: ICD-9 — 4330A\|4331A\|4339A\|4340A\|4341A\|4349A\|436<br>Hospital discharge: ICD-8 — 433\|434\|436<br>Hospital discharge: excluded ICD-10 — I63.6<br>Cause of death: ICD-10 — I63, I64<br>Cause of death: ICD-9 — 4330A\|4331A\|4339A\|4340A\|4341A\|4349A\|436<br>Cause of death: ICD-8 — 433\|434\|436<br>Cause of death: excluded ICD-10 — I63.6 |
| All Stroke | I9_SAH,<br>I9_ICH,<br>I9_STR_EXH,<br>I9_OTHINTRACRA | Hospital discharge: ICD-10 — I60,I61,I62,I63, I64<br>Hospital discharge: ICD-9 — 430\|431\|4330A\|4331A\|4339A\|4340A\|4341A\|4349A\|436<br>Hospital discharge: ICD-8 — 430\|431\|433\|434\|436<br>Hospital discharge: excluded ICD-10 — I63.6<br>Cause of death: ICD-10 —I60, I61,I62, I63, I64<br>Cause of death: ICD-9 — 430\|431\|4330A\|4331A\|4339A\|4340A\|4341A\|4349A\|436<br>Cause of death: ICD-8 — 430\|431\|433\|434\|436<br>Cause of death: excluded ICD-10 — I63.6 |
| COPD | J10_COPDNAS | Hospital discharge: ICD-10 — J44<br>Hospital discharge: ICD-9 — 4912<br>Hospital discharge: ICD-8 — 49104<br>Cause of death: ICD-10 — J44<br>Cause of death: ICD-9 — 4912<br>Cause of death: ICD-8 — 49104 |
| Chronic Liver Disease | K11_CHRONHEP,<br>CHIRHEP_NAS,<br>K11_ALCOLIV,<br>K11_FIBROCHIRLIV,<br>NAFLD | Hospital discharge: ICD-10 — K70,K73,K74,K76.0<br>Hospital discharge: ICD-9 — 5714A\|5714X\|571[0-3]\|571[5-6]<br>Hospital discharge: ICD-8 — 57001,5710,5719<br>Cause of death: ICD-10 — K70,K73,K74,K76.0<br>Cause of death: ICD-9 — 5714A\|5714X\|571[0-3]\|571[5-6]<br>Cause of death: ICD-8 — 57001,5710,5719 |

| | | |
|---|---|---|
| Chronic Kidney Disease | N14_CHRONKIDNEYDIS | Hospital discharge: ICD-10 — N18<br>Hospital discharge: ICD-9 — 585<br>Cause of death: ICD-10 — N18<br>Cause of death: ICD-9 — 585<br>KELA reimbursements: KELA codes — 138 |
| Alzheimer’s | G6_ALZHEIMER,<br>G6_NEURODEGENERATIVE | Hospital discharge: ICD-10 — F00,G30<br>Hospital discharge: ICD-9 — 3310<br>Hospital discharge: ICD-8 — 29010<br>Cause of death: ICD-10 — F00,G30<br>Cause of death: ICD-9 — 3310<br>Cause of death: ICD-8 — 29010<br>KELA reimbursements: KELA codes — 307<br>KELA reimbursements: ICD-10 — F00*, G30<br>Medicine purchases: ATC — N06D |
| Parkinson's and parkinsonism | G6_PARKINSON,<br>G6_PARKSCND | Hospital discharge: ICD-10 — G20,G21, G22*, G22*A52.1<br>Hospital discharge: ICD-9 — 3320A, 3321<br>Hospital discharge: ICD-8 — 34200,3420[8-9]<br>Cause of death: ICD-10 — G20,G21, G22*, G22*A52.1<br>Cause of death: ICD-9 — 3320A,3321<br>Cause of death: ICD-8 — 34200,3420[8-9]<br>KELA reimbursements: KELA codes — 110 |
| Rheumatoid Arthritis | M13_RHEUMA | Hospital discharge: ICD-10 — M05, M06<br>Hospital discharge: ICD-9 — 7140A\|7140B\|7241\|7142<br>Hospital discharge: ICD-8 — 712[1-3]<br>Cause of death: ICD-10 — M05, M06<br>Cause of death: ICD-9 — 7140A\|7140B\|7241\|7142<br>Cause of death: ICD-8 — 712[1-3] |
| Macular Degeneration | H7_MACULADEGEN | Hospital discharge: ICD-10 — H35.30, H35.31, H35.36#, H35.37#, H35.38#, H35.39#<br>Hospital discharge: ICD-9 — 3625<br>Hospital discharge: ICD-8 — 37710<br>Cause of death: ICD-10 — H35.30, H35.31, H35.36#, H35.37#, H35.38#, H35.39#<br>Cause of death: ICD-9 — 3625<br>Cause of death: ICD-8 — 37710 |
| Osteoporosis | M13_OSTEOPOROSIS | Hospital discharge: ICD-10 — M80, M81, M82*<br>Hospital discharge: ICD-9 — 733[0-1]<br>Hospital discharge: ICD-8 — 7230\|72391<br>Cause of death: ICD-10 — M80, M81, M82*<br>Cause of death: ICD-9 — 733[0-1]<br>Cause of death: ICD-8 — 7230\|72391 |
| Osteoarthritis | M13_ARTHTROSIS_COX,<br>M13_ARTHROSIS | Hospital discharge: ICD-10 — M15-M19<br>Hospital discharge: ICD-9 — 7151E\|7152E<br>Hospital discharge: ICD-8 — 71300<br>Cause of death: ICD-10 — M15-M19<br>Cause of death: ICD-9 — 7151E\|7152E<br>Cause of death: ICD-8 — 71300 |

**Table S3. LGBM hyperparameter search space**

| Parameter | Search Space |
|---|---|
| Boosting Type | GBDT |
| Learning Rate | (1e-3, 0.3) |
| Number of Estimators | (50, 500) |
| Number of Leaves | (15, 512) |
| Max Depth | (3, 12) |
| Minimum Data in Leaf | (5, 200) |
| Feature Fraction | (0.5, 1.0) |
| Bagging Fraction | (0.5, 1.0) |
| Bagging Frequency | (1, 10) |
| Lambda L1 | (1e-8, 10.0) |
| Lambda L2 | (1e-8, 10.0) |
| Min Gain to Split | (0.0, 1.0) |
| Scale Pos Weight | Num Controls / Num Cases |

**Table S4. MLP hyperparameter search space**

| Parameter | Search Space |
|---|---|
| Number of Layers | (3,6) |
| Dropout | (0.1,0.5) |
| Activation | (Relu, Selu, Elu) |
| Learning Rate | (1e-4,1e-2) |
| Units | (32,1024) |
| Batch size | (64,128,256,512) |

#### Table S5. FINDRISC criteria mapped to UKB fields

| FINDRISC question | Response / category | UKB Fields | Points |
|---|---|---|---|
| **1) Age** | **< 45 years** | 21022, Age at recruitment | 0 |
| | **45–54 years** | | 2 |
| | **55–64 years** | | 3 |
| | **≥ 65 years** | | 4 |
| **2) BMI** | **< 25 kg/m²** | 21002, Weight (kg)<br>50, Standing height (cm) | 0 |
| | **25 to <30 kg/m²** | | 1 |
| | **≥ 30 kg/m²** | | 3 |
| **3) Waist circumference** | **Men: <94 cm / Women: <80 cm** | 48, Waist circumference (cm) | 0 |
| | **Men: 94–102 cm / Women: 80–88 cm** | | 3 |
| | **Men: >102 cm / Women: >88 cm** | | 4 |
| **4) Physically active at least 30 min daily?** | **Yes** | If the sum of these is 7 days/week:<br>864, Number of days/week walked 10+ minutes<br>884, days/week moderate 10+ min<br>904, days/week vigorous 10+ min<br>AND the sum of these is >30min:<br>874, Duration of walks (min)<br>894, duration of moderate activity<br>914, duration of vigorous activity | 0 |
| | **No** | | 2 |
| **5) Eats vegetables / fruit (berries) every day?** | **Yes** | If the sum of these is at least 4 tbsp:<br>1289, Cooked vegetable intake (tbsps)<br>1299, Salad/raw vegetable intake (tbsps)<br>1309, Fresh fruit intake (tbsps)<br>1319, Dried fruit intake (tbsps) | 0 |
| | **No** | | 1 |
| **6) Ever taken antihypertensive (BP-lowering) medication regularly?** | **No** | | 0 |
| | **Yes** | 6153, Medication, Value 2 for blood pressure lowering medication for females<br>6177, same but for males | 2 |
| **7) Ever had high blood glucose?** (exam / illness / pregnancy, etc.) | **No** | | 0 |
| | **Yes** | 404, gestational diabetes only<br>Glucose measure (30740) > 7 if fasting time > 8 else if glucose >11 | 5 |
| **8) Family history of diabetes** | **No family history** | | 0 |
| | **Second-degree relative** (grandparent, aunt, uncle, first cousin) | Not available | 3 |
| | **First-degree relative** (parent, sibling, child) | Illness of father 20107, value 9 for diabetes<br>20110 for mother<br>20111 for siblings | 5 |

**Table S6. Average model performance across architectures**

| | L2 LogReg | L1 LogReg | MLP | LGBM | Transformer |
|---|---|---|---|---|---|
| UKB Prevalent | 0.8183 | 0.8175 | 0.7918 | 0.7875 | 0.8118 |
| UKB Incident | 0.7412 | 0.7338 | 0.7182 | 0.7227 | 0.7215 |
| FinnGen Prevalent | 0.8297 | 0.8311 | 0.7837 | 0.7713 | 0.8012 |
| FinnGen Incident | 0.7502 | 0.7504 | 0.7275 | 0.703 | 0.7333 |
| AoU Prevalent | 0.7831 | 0.7703 | 0.7637 | 0.7561 | 0.7732 |
| AoU Incident | 0.7521 | 0.743 | 0.7275 | 0.7281 | 0.7105 |

**Table S7. Comparison of all architectures to ridge regression**
We compare the AUROC for each architecture to the ridge regression across diseases using a paired t-test. We report the t statistic (positive indicating better performance in L2 than the compared model) and fdr adjusted p value.

| | L1 LogReg | MLP | LGBM | Transformer |
|---|---|---|---|---|
| UKB Prevalent | t=0.19, p = 0.89 | t=2.73, p=0.051 | t=2.62, p=0.051 | t=1.54, p=0.18 |
| UKB Incident | t=2.25, p=0.089 | t=4.29, p=0.0088 | t=1.94, p=0.072 | t=1.68, p=0.17 |
| FinnGen Prevalent | t=-0.28, p=0.85 | t=3.10, p=0.042 | t=2.71, p=0.051 | t=1.49, p=0.18 |
| FinnGen Incident | t=-0.024, p=0.98 | t=2.10, p=0.11 | t=4.98, p=0.0052 | t=1.57, p=0.17 |
| AoU Prevalent | t=1.61, p=0.17 | t=2.64, p=0.051 | t=1.71, p=0.17 | t=1.88, p=0.14 |
| AoU Incident | t=1.59, p=0.17 | t=2.73, p = 0.051 | t=3.04, p=0.042 | t=3.42, p = 0.033 |

**Table S8. Predictive power of top 20 PCs vs proteomics data**

| Disease | PC Prevalent | Proteomic Prevalent | PC Incident | Proteomic Incident |
|---|---|---|---|---|
| Type 2 diabetes | 0.8435 | 0.9605 | 0.7454 | 0.8925 |
| Chronic liver disease | 0.7792 | 0.9124 | 0.7623 | 0.8544 |
| COPD | 0.7772 | 0.8501 | 0.7834 | 0.8519 |
| Chronic kidney disease | 0.8177 | 0.8472 | 0.813 | 0.8424 |
| Alzheimer's disease | | | 0.6953 | 0.8384 |
| All cause stroke | 0.6762 | 0.7907 | 0.6995 | 0.7882 |
| Ischemic Stroke | 0.6614 | 0.7643 | 0.6888 | 0.7738 |
| Parkinson's disease | 0.5459 | 0.9466 | 0.6885 | 0.7411 |
| Osteoporosis | 0.7546 | 0.8585 | 0.6511 | 0.7337 |
| Osteoarthritis | 0.7076 | 0.7637 | 0.6181 | 0.6887 |
| Ischemic heart disease | 0.7097 | 0.7883 | 0.6456 | 0.6813 |
| Rheumatoid arthritis | 0.6843 | 0.7996 | 0.5248 | 0.6538 |
| Macular degeneration | 0.5777 | 0.7097 | 0.6118 | 0.6443 |
| Asthma | 0.5654 | 0.6834 | 0.4769 | 0.5696 |
| Ulcerative colitis | 0.5575 | 0.7805 | 0.4183 | 0.5634 |
